# Economic Loss due to Health Funding Cuts as Distributed Across Geospatial Units

**DOI:** 10.1101/2025.07.24.25332092

**Authors:** Mallory J. Harris, Alyssa H. Sinclair, Elena K. Parkerson, Clio Andris, Joshua S. Weitz

## Abstract

In 2025, the National Institutes of Health (NIH) proposed a 15% cap on indirect research costs and the Department of Health and Human Services (HHS) terminated or froze over 5,000 grants. These policy changes would lead to billions of dollars in annual future economic losses and jeopardize billions of dollars in ongoing economic activity associated with medical research. Assessing the local economic impacts of cuts to NIH research requires accounting for commuting flows between where individuals work and live rather than assuming that impacts are confined to affected institutions. We estimate that 60% of U.S. counties will experience over $100,000 in economic losses from the indirect cost caps if they are implemented – far more widespread than the 12% of counties where NIH-supported grantees are located. Likewise under a commuter model, 22% of counties could lose more than $100,000 due to NIH grant terminations and freezes, again more widespread than the 4% of counties containing affected institutions. Incorporating commuting flows and accounting for economic multipliers of research funding reveals the extent to which biomedical research funding cuts will lead to substantial economic losses in local communities throughout the U.S.

## 1 Introduction

Scientific research spending drives economic activity by creating and maintaining jobs, supporting purchases of research-related material, and increasing productivity [40, 2, 47, 12]. Cuts to research may therefore lead to economic losses in the form of lost tax revenue, purchasing power, and salaries [8]. On February 7, 2025, the NIH introduced a policy to substantially reduce funding for indirect costs (IDCs) of research [30]. IDCs represent the costs associated with grant administration, safety compliance, contract negotiation, equipment, cleaning and maintenance, utilities, and other operations and materials that support science. The proposed policy would cap indirect costs (IDCs) at 15% of the direct costs associated with each grant [30], a significant reduction from current levels — currently, realized indirect costs are estimated to be *∼* 42% of direct costs [2]. More than 5,800 research grants totaling nearly $9.0B in research support had been terminated or frozen by the U.S. Department of Health and Human Services (HHS) between February and December of 2025. According to White House statements and termination letters, grants have been terminated because they no longer align with revised agency priorities (e.g., many targeted grants included a focus on climate change, health equity, COVID, and vaccines) and/or grant recipients are affiliated with certain, targeted universities (e.g., Harvard University and Columbia University) [24, 33, 32]. However, the local economic impacts on communities arising from cuts to federal support of medical research remains underexplored.

Efforts to measure and communicate the local economic impacts of cuts to biomedical research can build public awareness and motivate action-taking [35, 36, 5]. In doing so, analyses focused on the impacts of these cuts at an institutional level help articulate what is lost [27, 28, 3, 2]). Many universities have released public communications detailing the consequences of cuts to research at their institutions [37, 32, 38, 46, 6] while others have remained largely silent. Although prior analyses and communications highlight harm to specific institutions, an institution-centered approach obscures the extent to which negative economic impacts extend beyond the immediate geographic vicinity of research institutions. This spatially-extended impact arises, in part, because institutions with NIH grants are often major employers of commuters and purchasers of products from surrounding areas, meaning that those areas are likely to experience declines in tax revenue and spending due to research funding cuts. Here, we analyze public data on NIH grants to reveal new policy-relevant insights. We estimate the economic impact of cuts to funding for IDCs and terminated grants and demonstrate that integrating quantitative data on employee movement geographies (i.e., commutes) provides a more comprehensive near-term assessment of the economic impacts of biomedical research cuts on local communities throughout the United States. To quantify this impact, we model the spatial distribution of future economic losses due to IDC funding cuts and current economic losses due to terminated/frozen grants across jurisdictions using commuter flows from the U.S. Census from counties to tracts. We demonstrate that accounting for commuter flows reveals broader and more diverse community impacts than estimates produced by prevailing methods of mapping losses that use only the location of the institution, without considering workers’ home locales. Focusing on connected geospatial units reveals the widespread negative impact of NIH funding cuts and we recommend that efforts to map and aggregate data illustrating current and future losses account for commuter flows.

## 2 Methods

### 2.1 Data and processing

#### 2.1.1 NIH Grant Data for IDC Reduction

Given disruptions to awarding of new grants in fiscal year 2025, we estimated the potential impact of a future cap on indirect costs based on grants awarded in fiscal year 2024, assuming that future awards will be distributed geographically in an approximately similar manner. NIH data were retrieved from NIH RePORTER (https://reporter.nih.gov/) for all grants that were active in fiscal year 2024, totaling 78,069 entries. For each award, available data include direct costs, indirect costs, awardee organization name, and awardee coordinates (latitude and longitude). We excluded 12,324 entries without a Catalog of Federal Domestic Assistance (CFDA) code, as these entries indicate sub-awards (e.g., cores within a center grant) when total funds are already accounted for in the center grant. The final dataset included 65,745 grants. In two cases where a total grant value was listed without division into direct and indirect costs, we assumed no indirect costs.

#### 2.1.2 Data on Terminated/Frozen Grants

We obtained data on reported NIH grant terminations and freezes as of December 26, 2025 from Grant Witness, a publicly available dataset [34]. The Grant Witness database is updated regularly based on integrated data from news reports, the HHS TAGGS system, DOGE.gov, USAspending.gov, NIH’s account on X, and NIH RePORTER, as well as self-reporting by scientists. Grants that were terminated or frozen but subsequently reinstated by the time of analysis were excluded from loss calculations.

#### 2.1.3 Census Unit Assignment

We mapped grant data at the coordinate (latitude, longitude) level based on coordinates provided in the NIH RePORTER data. Although satellite campuses and teaching hospitals located some distance from the main campus are sometimes disaggregated (e.g., University of Miami main campus versus University of Miami School of Medicine; The University of Texas at San Antonio Health Science Center versus The University of Texas San Antonio), the coordinates provided are often administrative buildings rather than particular research facilities. Some institutions may have research facilities spread throughout the region or conduct projects in areas that are distant from the institution. As a sensitivity analysis, we tested the impacts of geolocating two large multi-campus institutions (UCLA and Johns Hopkins University) to each of their affiliated medical locations (329 and 91 locations respectively). We joined point-level records to census tracts, congressional districts, U.S. county TIGER/Line shape files using a spatial join. Block group centroids that did not overlap with the congressional district polygons (e.g., whose centroid is on the coast) were assigned to their nearest congressional district. One record did not have valid coordinates and was therefore excluded. By comparing parcel data to census tract assignments across five states, we confirmed that 564/566 institutions that could be tested were contained by their assigned tracts using a majority-of parcel criterion (21 additional institutions in New York could not be tested due to incompleteness of the state parcel dataset) (Supplemental File 1).

#### 2.1.4 Classifying Regions Based on Urbanicity and Partisanship

We classified counties as metropolitan (metro) or non-metro based on 2023 Rural-Urban Continuum Codes provided by the U.S. Department of Agriculture (USDA) (https://www.ers.usda.gov/data-products/rural-urban-continuum-codes/documentation) [20]. U.S. House of Representatives congressional districts were classified as Democratic or Republican (or Other) based on the party affiliation of their representative elected to the 119th Congress. In a supplemental analysis, impacts were summarized according to categories defined by the American Community Project based on socioeconomic and political characteristics [21].

#### 2.1.5 Commuter Flows

We used *commuter flows* for all job types (JT00) from the Origin-Destination Employment Statistics (LODES)-Longitudinal Employer Household Dynamics (LEHD) data set (version LODES8) provided by the U.S. Census [42]. These data are released at the census block level (JT00 2016). We used the 2016 dataset, as this is the most recent dataset that includes all states (Alaska has not had full coverage since 2016).

We summed flows from census blocks to produce area-to-tract flows, tracing commuters from home areas (either counties or congressional districts) to tracts where workplaces are located. We created centroids (center points) of the block groups and spatially joined the centroids with congressional district polygon files (500K scale) for the 119th Congress (Jan 2025–Jan 2027). We included all 435 districts with voting members, plus the district corresponding to Washington, D.C., which has a non-voting delegate.

### 2.2 Methods of estimating losses

#### Proposed indirect cost cap

We calculated losses from the proposed indirect cost cap for each grant by taking the difference between the listed indirect costs and the maximum indirect costs under the proposed policy (15% of the listed direct costs). In cases where this value is zero or negative (i.e., current indirect costs *<*15%), we assume no losses would be incurred. In other words:

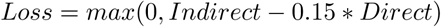

#### Terminated/frozen grants

All remaining funding from terminated/frozen grants (calculated by Grant Witness as total obligated funds minus outlays (i.e., disbursed funds) according to USASpending.gov) is treated as a loss. For a subset of multi-year supplements and subawards, only the budget for the current year is reported, leading us to underestimate the actual amount of funding lost [34].

#### Calculating economic loss

We estimate economic loss by applying a statelevel multiplier to IDC and terminated/frozen grant losses based on a report by United For Medical Research [40], which lists NIH spending, economic activity generated from NIH spending, and jobs supported from NIH spending by each state in FY2024. We estimate the ratio between economic activity generated or jobs created and NIH spending to determine the state-level economic and job multipliers respectively, which should be interpreted as short-term impacts. Estimated economic losses therefore correspond to outputs removed at the place of production. In a supplemental analysis, we estimated losses using a national multiplier of $2.56 derived from the same report.

### 2.3 Distributing losses based on commuter flows

We redistribute losses across spatial units based on commuter flows. For each destination tract, we estimate the total number of people who work in a given spatial unit A by summing total commutes with unit A as the destination. For each spatial unit B (county or congressional district) that houses residents who work in unit A, we then calculate the proportion *p_A,B_* of people working in tract A who live in unit B. Unit B will experience losses from tract A (*L_A_*) proportionally to its share of commuters, *p_A,B_ ∗ L_A_* (this process is visualized across a set of example counties in Figure 1). Total loss to unit B is then the sum of its share of losses across all impacted tracts to which its residents commute, including the contributions from losses in unit B (as applicable) that are not redistributed.

**Figure 1:**
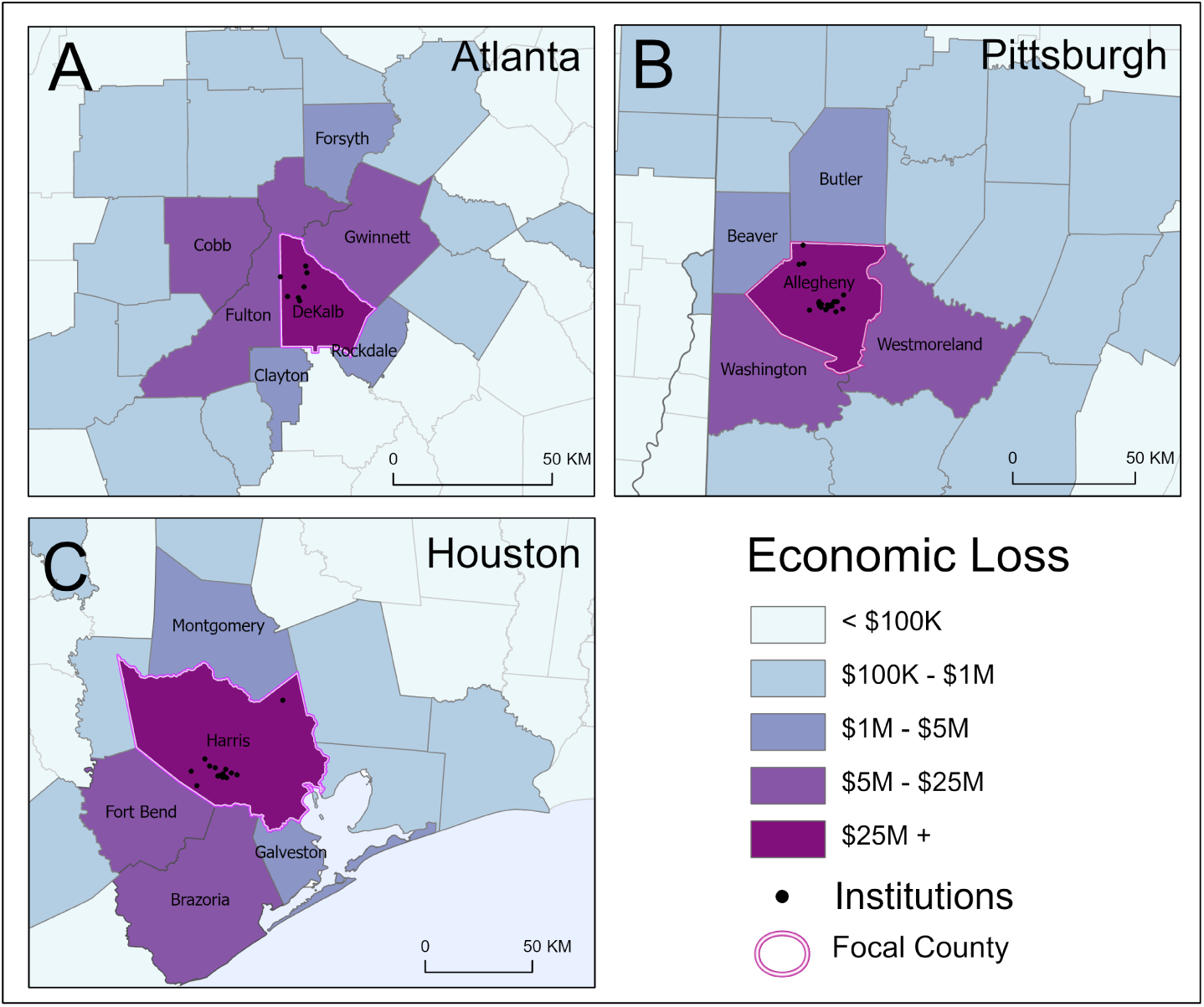
Economic losses extend outward from the counties where they originate. Highlighted counties include major cities: (A) Dekalb County, Georgia (Atlanta); (B) Allegheny County, Pennsylvania (Pittsburgh); and (C) Harris County, Texas (Houston). Counties are colored by future estimated annual economic loss due to the proposed cap on indirect costs based on commuter flows that connect to a focal county (indicated by light pink outline). Institutions within focal counties are indicated as black points. State outlines are indicated with darker lines.

In a sensitivity analysis, we defined a parameter *ρ* as the proportion of funding that is treated as static (i.e., remains in the tract where an institution is located); the remaining fraction of the funding (1 *− ρ*) was redistributed under the commuter model. We tested the effects of varying *ρ* from one (static model) to zero (commuter model). In another sensitivity analysis, we filter commutes to sectors relevant to indirect costs. We use Residence Area Characteristics (RAC) and Workplace Area Characteristics (WAC) files from the U.S. Census for the most recent year available at the tract level (2016) [42], which provide the total number of residents or workers based on North American Industry Classification System (NAICS) two-digit sector codes. We then rescaled the total commuters following a given flow by the geometric mean of the proportion of residents in relevant sectors commuting from the origin county and the proportion of workers in relevant sectors commuting to the destination tract. We then distributed losses as above based on the rescaled commuter numbers. Given that there is some uncertainty in exactly which sectors could be impacted by NIH funding cuts, we conduct the analysis using two different sets of relevant sectors: one broader and one more narrow. The broad set of sectors is: Utilities (CNS03), Construction (CNS04), Manufacturing (CNS05), Retail Trade (CNS07), Transportation and Warehousing (CNS08), Professional, Scientific, and Technical Services (CNS12), Management of Companies and Enterprises (CNS13), Admin/Support & Waste Management (CNS14), Health Care and Social Assistance (CNS16). The narrow set of sectors is CNS12, CNS13, CNS14, and CNS16.

## 3 Results

We calculate anticipated annual losses associated with the IDC cap at the county level (*n*=3, 144 units) with a static model (i.e., without accounting for commuter flows) versus with a commuter model (as illustrated in Figure 1). In a static model, the majority of counties do not register any loss (88% of counties or 2,768 counties with no losses from the IDC cap) and losses are concentrated in major metropolitan areas (Figure 2A). The greatest economic losses are in Suffolk County, Massachusetts ($1.0 billion) and New York County, New York ($1.0 billion) (including Boston and Manhattan, respectively). Without applying commuter flows, we estimate a total of $15.7 billion in economic losses from the proposed indirect cost cap, applied to institutions located in 12% of counties (376 counties).

**Figure 2:**
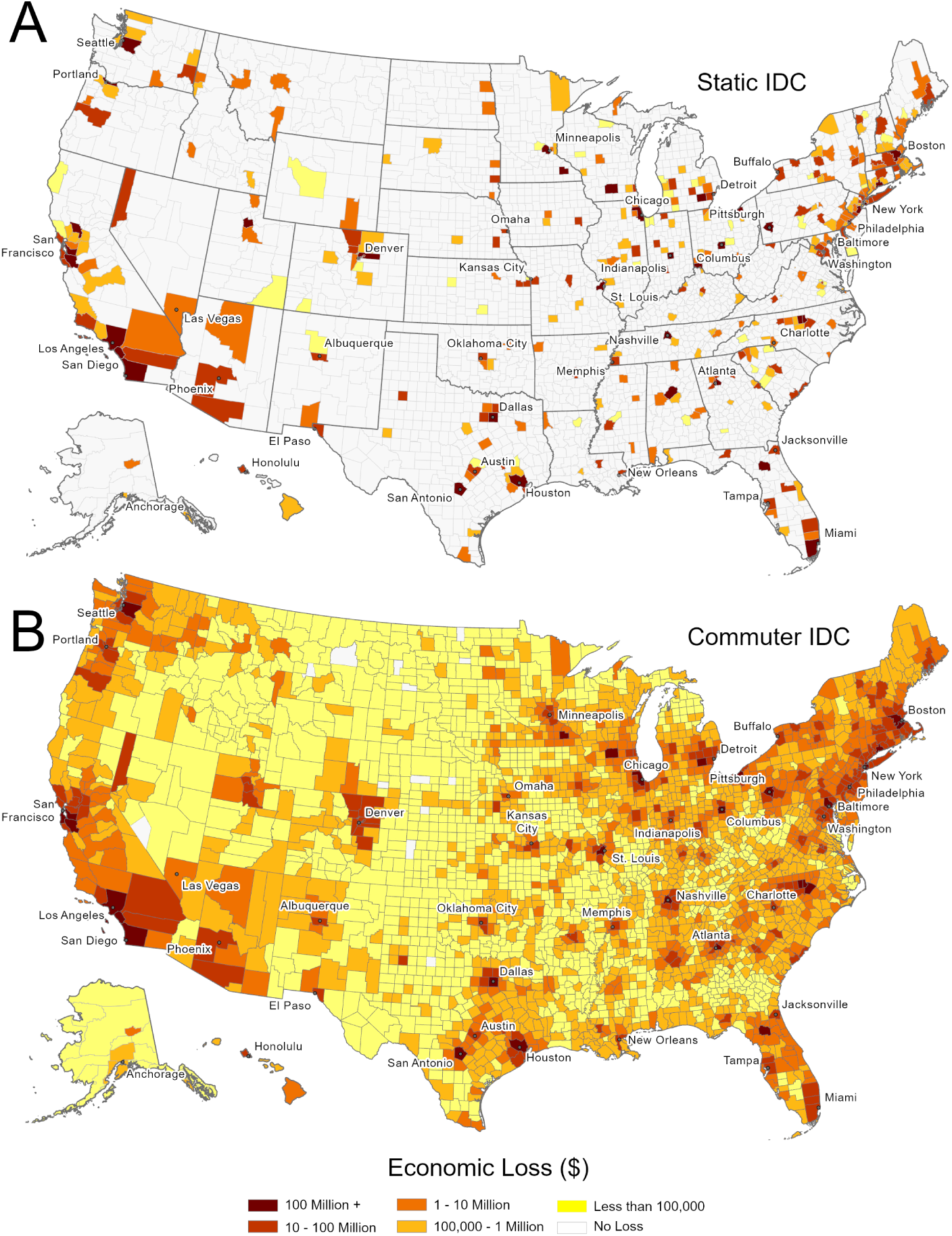
Economic losses from the proposed IDC cap are contained to a subset of counties when estimated using a static model (A), while losses are widespread when using the commuter model (B). Maps indicate economic losses (in USD) across the United States at the county level. County-level values are available and archived on Zenodo at doi.org/10.5281/zenodo.22833910.

In contrast, in the commuter model the $15.7 billion in economic losses impacts nearly all 3,144 counties and 60% of counties (1,873 counties) incur more than $100,000 in economic losses annually (1,499 or 80% of which do not experience any losses in the static model) (Figure 2B). Without accounting for economic multipliers, under the commuter model 40% of counties (1,281 counties) incur more than $100,000 in projected raw (unmultiplied) losses annually (Figure S1A). All congressional districts are expected to incur more than $100,000 in economic losses (Figure S2). In total, $7.1 billion (constituting 45% of all losses) would be reapportioned to counties other than that associated with grantee institutions. Counties with no estimated losses under the static model incur considerable economic losses under the commuter model (mean: $750,032 and median: $117,987) (Figure 3A). In counties with non-zero estimated losses under the static model, estimate losses under the static and commuter model are significantly correlated (*ρ* = 0.74, *p <* 0.001, Figure 3B).

**Figure 3:**
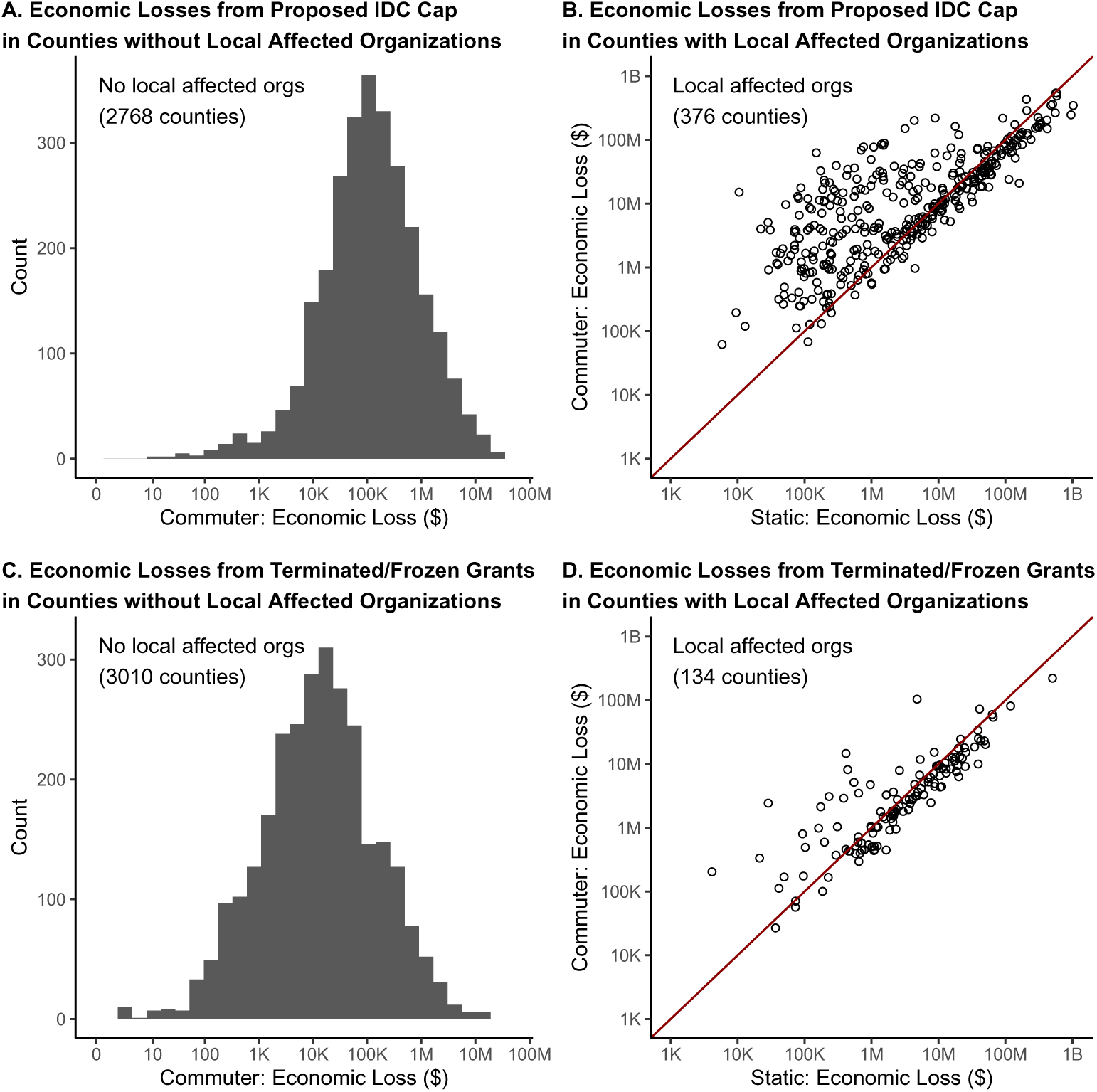
Across most counties, accounting for commuter flows increases the estimated economic losses from NIH funding cuts. Faceted plots compare the impacts of economic losses from the proposed cap on indirect costs (top, A and B) and terminated grants (bottom, C and D). Histograms (left, A and C) indicate losses under the commuter model across counties with no local affected organizations (i.e., no losses under the static model). Scatterplots (right, B and D) compare losses under the commuter versus static model in counties with local affected organizations (e.g., non-zero losses under the static model). The y = x line is shown in red, with points above the line corresponding to counties with greater losses under the commuter model compared to the static model. In all panels, annotations indicate the number of counties meeting the corresponding criteria. Economic losses are displayed in log-scaled USD.

Some counties’ estimated losses change by orders of magnitude when accounting for commuter flows (Figure 3). The following areas have a relatively high share of *outgoing* economic impacts of funding cuts when accounting for commuter flows: Adams County, CO (home to the University of Colorado Anschutz Medical Campus); Charlottesville, VA (home to the University of Virginia); Montour Co., PA (home to the Geisinger Medical Center); New York County, NY; and Richmond, VA (home to Virginia Commonwealth University). Discounting commuter flows will overestimate the relative impact of NIH funding cuts in those focal counties. The following areas have a relatively high relative share of *incoming* economic impacts of funding cuts when accounting for commuter flows (Table 1): Arapahoe County, CO (near Denver); Brazoria County, TX (near Houston, Figure 1B); Gwinnett County, GA (near Atlanta, Figure 1A); Williamson County, TN (near Nashville); and Anne Arundel County, MD (near Washington, DC and Baltimore). These locales would not be recognized as losing notable (or any) funding without the commuter model. For both the county and district level estimates, the impact of research funding cuts on non-metropolitan areas and congressional districts represented by Republicans are more evident when using the commuter model (Figure S3, Table S1). There are 1,903 non-metropolitan counties and 55 Republican congressional districts (compared to 865 metropolitan counties and 23 Democratic congressional districts) that only experience economic losses when accounting for commuters. For counties and districts that have at least one organization funded by NIH and impacted by the IDC cuts, the ratio of losses under the static versus commuter model is approximately equivalent for non-metropolitan and metropolitan counties (non-metro median: 1.5, IQR: 0.8–6.5; metro median: 1.6, IQR: 0.8–10.5) but generally greater in Republican versus Democratic congressional districts (Republican median: 4.3, IQR: 1.1–47.5; Democratic median: 2.4, IQR: 0.7–22.3). Under a commuter model versus a static model, 3.5 times more metro counties and nearly 22.4 times more non-metro counties are projected to experience over $100K in economic losses from the IDC cap. Economic losses per 100,000 people tends to be greater across metropolitan ($5.1M versus $1.2M in non-metropolitan) and Democratic ($6.9M versus $2.5M in Republican districts). Similarly, across most types defined by the American Communities Project (e.g., exurbs, rural middle America, and military posts), a small number of counties are expected to experience over $100K in economic losses under a static model but almost all counties are expected to lose over $100K under a commuter model (Table S2). Anticipated economic losses tend to be greater in counties with a greater GDP (Figure S4). In sum, economic losses in non-metro counties and districts presently represented by a Republican are more apparent when accounting for commuter flows.

**Table 1:**
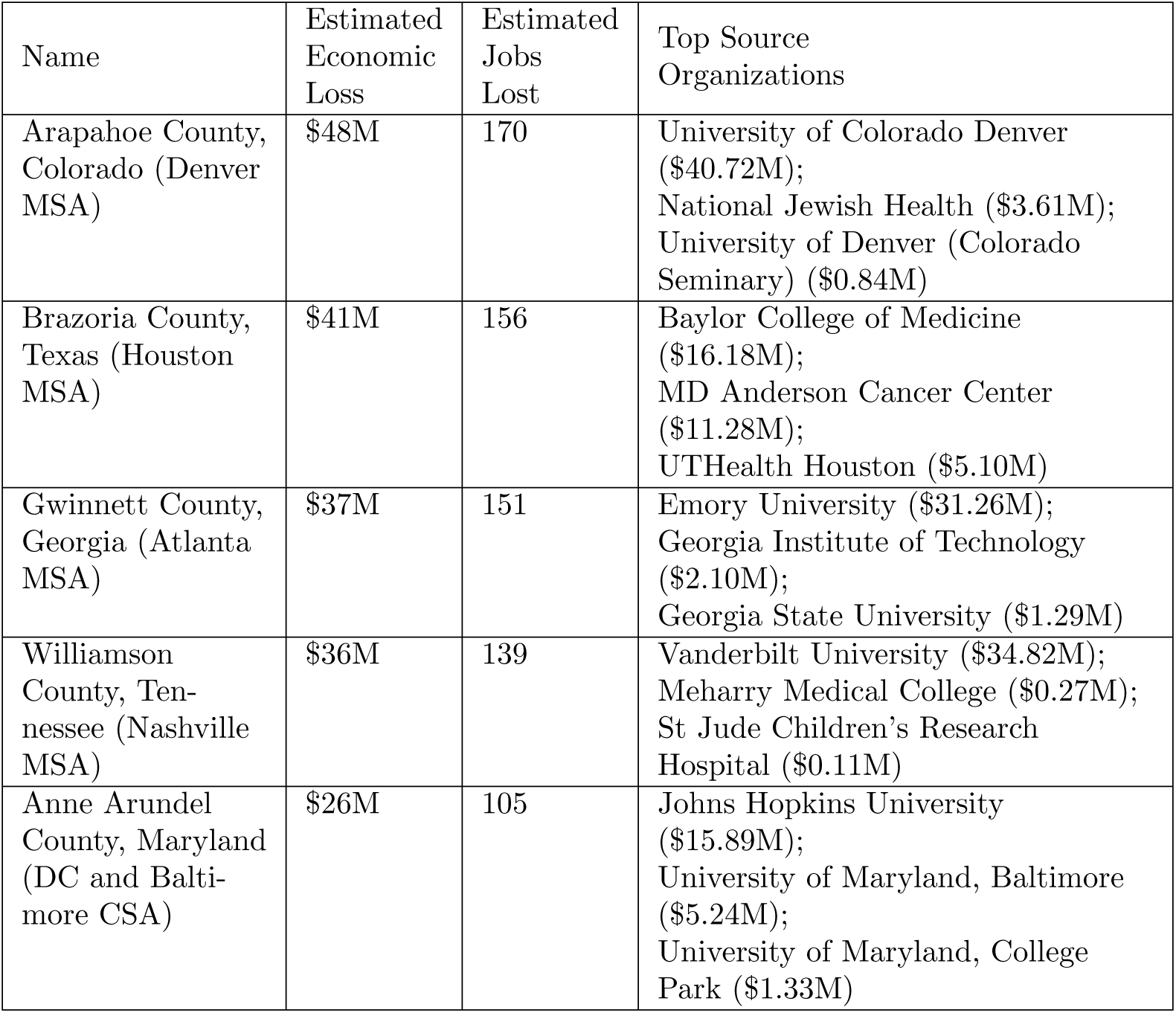
Several counties with no losses under the static model experience substantial losses under the commuter model. Five counties with the greatest economic losses from the proposed IDC cap under the commuter model and no losses under the static model are listed. The columns indicate economic losses under the commuter model (in millions of dollars), number of jobs lost under the commuter model, and the top three (proximal) organizations contributing to economic losses (with associated economic losses in millions of dollars in parentheses). MSA stands for metropolitan statistical area, CSA stands for combined statistical area.

We find similar results when examining the economic impact of *grant terminations and freezes*, i.e., losses incurred from canceled and frozen grants. We find that the total losses are widespread, significant in magnitude, and impact far more counties in a commuter model relative to estimates derived from a static model (Figure 3C, D). Specifically, as of December 26, 2025, we estimate $1.7 billion in economic losses are associated with grant terminations across institutions in 4% of counties (134 counties). The greatest economic losses are located in Durham County, North Carolina ($505M), Galveston County, Texas ($119M) and Cook County, Illinois ($65M). When commuter flows are applied, 22% of counties (680 counties) face economic losses of over $100,000 in NIH funding due to grant terminations (Figure 4, Figure S5). Moreover in the commuter model, a total of $825 million in economic losses (48% of losses) impact counties outside the county where an organization’s grants were terminated. Figure S5 shows comparisons of economic losses from terminated grants at the level of congressional districts. Under the commuter model, raw (unmultiplied) losses from terminated grants exceed $100K in 14% of counties (445 counties) (Figure S1B). The proposed cap on indirect costs affects a broader set of grants and has a larger total projected economic impact than realized grant terminations. Economic losses are robust to varying the share of funding that remains static (*ρ*) versus flowing based on commuting (Figure S6); over 1,000 counties are projected to lose more than $100,000 due to the IDC cap as long as at least 20% of funding follows commuter flows (*ρ ≤* 0.80) (Figure S7). Economic losses from the IDC cap and terminated/frozen grants are consistent when *ρ* = 0 compared to *ρ* = 0.25 (*R*^2^ = 0.83 and *R*^2^ = 0.90 respectively). The economic multiplier varies across states from 1.94 in Maryland to 6.10 in Nevada compared to the national average of 2.56 (Figure S8), but estimates are also robust when a nationwide economic multiplier is applied (*R*^2^ = 0.99, see Figure S9). Total estimated losses nationally from the IDC cap and terminated grants is consistent with the main analysis ($15.7B and $1.7B).

**Figure 4:**
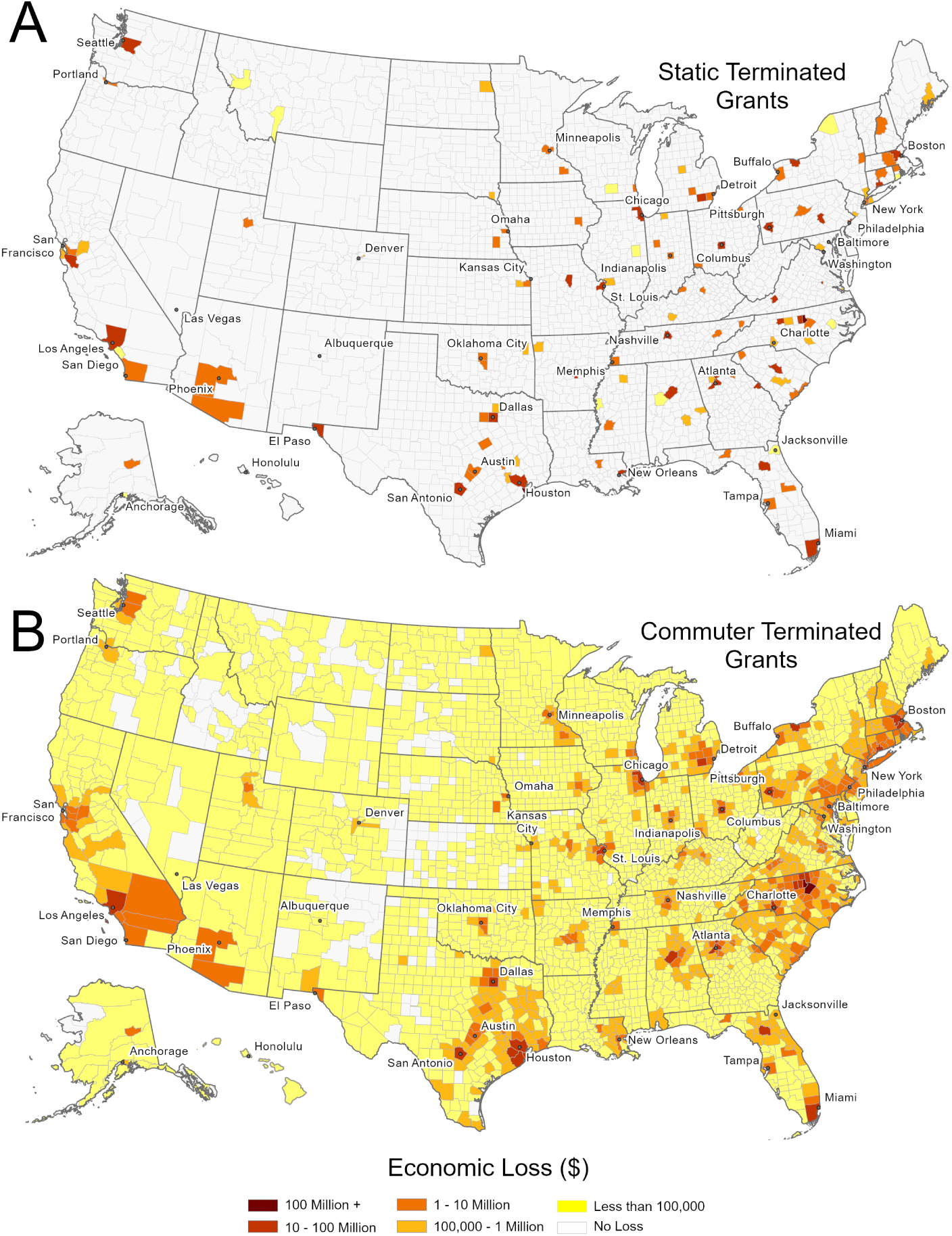
Economic losses from terminated grants are contained to a subset of counties when estimated using a static model (A), while losses are widespread when using the commuter model (B). Maps indicate economic losses (in USD) across the United States at the county level. County-level values are available and archived on Zenodo at doi.org/10.5281/zenodo.22833910.

Estimates are also robust to using the most recent commuter flows, with 60% and 22% of counties respectively expected to lose over $100K from the proposed IDC cap and terminated/frozen grants respectively (1,873 and 704 counties respectively) (*R*^2^ = 0.99 for economic losses from IDC cap, Spearman’s *ρ* = 0.94 and 0.93 for IDC cap and terminated/frozen grants across all 3,144 counties respectively, see Figure S11 and Figure S10). The proportion of counties with losses exceeding $100K is highly robust to the use of more recent commuter flows, with 5% of counties (177 counties) only exceeding this threshold for losses from the IDC cap when the analysis is conducted using more recent commuter flows (predominantly located in Missouri, Texas, Illinois, and Tennessee); an equal number of counties only exceed this threshold if 2016 flows are used (predominantly located in Georgia and Iowa). Similarly, 22 counties only have nonzero losses from the IDC cap under one set of commuter flows (14 counties with nonzero losses using most recent flows and eight counties with nonzero losses using 2016 flows) and 156 counties only have nonzero losses from terminated/frozen grants under one set of commuter flows (51 counties with nonzero losses using most recent flows and 105 counties with nonzero losses using 2016 flows). There is no shift in the number of metro/non-metro counties that exceed the $100K threshold for losses depending on the recency of commuter flows. In sum, the overall spatial pattern and county ranking of losses is robust to the reference year of commuter flows used, although individual county point estimates may vary. Results are also robust to filtering commuter flows to relevant sectors (*R*^2^ = 0.98 for both broad and narrow sectoral filtering, Figure S12), although nine counties have economic losses only without sectoral filtering (indicating that the residents who work in tracts containing affected institutions are not employed in sectors likely to be impacted by research funding cuts). The median economic losses from the IDC cap at UCLA and Johns Hopkins is generally consistent if grants are geolocated at satellite medical facilities where research may be conducted compared to geolocating grants to the main campus, but county-level estimates may be sensitive to the actual unreported distribution of funding across locations as there can be considerable variation in commuting connections between surrounding home counties and the census tracts containing satellite medical facilities (Figure S13 and Figure S14).

## 4 Discussion and Conclusion

In this work, we analyzed public data on NIH grants to generate policy-relevant insights about economic impact. We demonstrated the importance of accounting for commuter flows when assessing the economic impact of health research funding cuts across different geographic locations. Although a static model suggests that cuts will only affect specific institutions within a small subset of counties, a commuter-based model suggests that economic losses would be widespread throughout the country, affecting a broader and more diverse range of counties. These results illustrate the impact of NIH funding cuts on communities nationwide. To give a sense of scale, losses from the IDC cap is 0.015% of county-level GDP as the median percentage across counties, and the greatest percentage across counties is 1.4% in Hancock County, ME (Figure S4). An interactive map of our results under the commuter model is available at https://scienceimpacts.org/.

There are caveats associated with this approach. For example, the geographic level (e.g., county or congressional district level) to which data is aggregated will impact the specific quantitative degree of impact. This issue is known as the Modifiable Areal Unit Problem (MAUP) [13] and can be exacerbated when using flow data, as the aggregation affects both origin and destination [25, 45]. Estimates may be especially sensitive to a subset of institutions that are disproportionately impacted by the research funding cuts (e.g., those that have been targeted for grant disruptions and repeatedly had funding restored and disrupted through ongoing litigation processes). Other potential limitations of the model are related to the types of workers reflected in commuting flows and accuracy and recency of data. We used U.S. Census data that represents commuting flows from 2016, as more recent data exclude two states. Although commuter patterns may have changed in intervening years, our findings are robust to using more recent commuter data where available (Figure S11, Figure S10). These commuter flow data span all professions and workplaces, without accounting for commuting patterns that might be specific to research institutions or the impact of student workers (although our findings are robust to filtering commuter flows to sectors that are most likely to be affected by research funding cuts, see Figure S12). The multipliers used here reflect a range of economic benefits of spending, including interindustry spending on goods used at the research institution and increased spending by households due to income connected to research. Some proportion of economic losses may therefore remain local to a given institution rather than following commuter flows. As a relevant estimate, two large academic research organizations and the National Institute of Allergy and Infectious Diseases (NIAID) budget guide report that 50%-80% of NIH grant spending goes to personnel [41, 44, 29] and therefore a substantial share of economic impacts likely reflect commuter patterns. We project widespread economic losses in most counties in the United States throughout this range (Figure S7). Institutions were geolocated to census tracts based on coordinates provided to NIH. Losses to distant firms and employees who sell medical materials and equipment are not captured in this model, as trade (and lack thereof) is not considered (although vendors may be located at a substantial geographic distance from the institution when specialized equipment is required, see Weinberg et al. [47]). The model also does not account for research conducted at affiliated medical facilities that may be spread throughout a region (Figure S13 and Figure S14), collaborative grants that may span institutions, and geographic spillover of economic benefits beyond commuter patterns. Together, these two omissions likely bias our analysis toward underestimating the true economic impacts of reduced NIH funding outside of urban centers with large universities, although we still detect substantial losses across such communities.

Estimates of economic impact rely on local economic multipliers applied to funding lost from proposed cuts to research infrastructure and past and ongoing grant terminations. The United for Medical Research [40] report state-specific economic multipliers for NIH research spending have several limitations relevant to this analysis. The Regional Input-Output Modeling System II (RIMS II) economic multiplier estimates rely on parameters from the Bureau of Economic Analysis for calendar year 2022 due to lags in the economic data [4]. Some impacts of research funding that are included in the economic multiplier estimate (e.g., revenues connected to intellectual property) may not be relevant to each type of cut. A key limitation is that the multiplier is state-specific whereas markets and commuter flows are not bound to states. Due to interstate commuting, a small percentage of losses (5% of raw losses from the proposed cap on indirect costs) are redistributed outside of the state where the affected institution is located while using the state multiplier associated with the affected institution. A benefit of this approach is that it allows for a more realistic calculation of the local impact of losses, as dollar values are not ‘felt evenly’ across regions that vary in household purchasing power, real estate and land value, and institutional value. Further, economic impacts of research funding cuts likely also depend on the existing resources of the institution and its surrounding community, as well as the particular types of research and the type of job lost (e.g., career stage). The economic impact of NIH spending varies by state, tending to be greater in more rural, less populous states (Figure S8). Impacts of realized cuts could be measured directly across heterogeneous local contexts to characterize such differences at a more granular geographic scale. We rely on economic multipliers derived for all NIH research spending as a proxy for the impacts of marginal reductions in funding. Amid reports that universities are already reducing budgets due to proposed and realized federal funding cuts, additional efforts may apply causal methods (e.g., event study frameworks) to detect and quantify economic losses that have already occurred [22] and revisit the impacts of historical fluctuations in NIH funding.

This research is aligned with but does not take full advantage of equilibrium models and dynamic models of labor that estimate economic impacts of worker and capital movement across geographic space. Such models help estimate regional asymmetries associated with attracting labor and push and pull factors of workers due to conditions such as higher wages in another destination [9, 17, 39], or auxiliary factors such as better public transportation [14], ‘richness’ of amenities (such as museums, restaurants, libraries, schools) [18], temperature differentials [10], and gender balances [7]. Similar models that are relevant to our discussion posit that denser environments with ‘knowledge-intensive services’ and scientific innovation have significant pull factors [26] and that firm agglomeration through product variety enhances labor market competitiveness [15]. While these models require greater consideration of feedback within dynamic economic systems and may necessitate local and national labor market conditions estimation of several parameters with geographic variation, our geographically-weighted network model is flexible and can be applied to heterogeneous local economic contexts ranging from small college towns to large metropolitan areas. The model can be used to estimate losses rapidly and transparently in response to funding policy changes. In addition, a reliable migration model that captures fluctuation in cuts and threats to cuts requires data with a longer time series than we had access to in this study. Future work could measure longer-term effects, including how workers migrate due to job loss as a result of science cuts, as well as their re-employability both locally and in distant labor markets. Practically, some areas experiencing layoffs due to science cuts (e.g., Los Angeles) could also provide other jobs for those impacted where other areas (e.g., small college towns) may not.

Tracking research funding cuts is particularly difficult in light of rapid changes that are not clearly or reliably documented by federal sources. Our evaluation of terminated and frozen grants therefore relies on a crowd-sourced database that integrates reports of terminations/freezes from multiple sources including the NIH, HHS, and self-reports; these records may not consistently align with federal reports [34]. Scientific funding cuts have been the subject of multiple lawsuits, which may lead to complete or partial restoration of funding. In January 2026, an appeals court upheld the ruling that changes to the indirect cost rate must be made by Congress, a point reaffirmed by Congress in the FY2026 appropriations bill [1]. An estimated $10B in economic losses from terminated or frozen grants have been averted since August 2025 due to grant restorations [19]. On June 16, 2025, a federal judge ruled that terminated grants should be restored in a case brought by the Democratic attorneys general of sixteen states. However, only terminated grants listed by the plaintiffs were addressed, suggesting that impacts may become increasingly geographically heterogeneous, potentially along partisan lines [31] – even as grant terminations have continued. While we present a snapshot of current impacts from disrupted grants at a particular point in time, daily-updating estimates are available at https://scienceimpacts.org/. All code and data used to produce these analyses are published, which we hope may facilitate complementary analyses of additional scenarios.

Here, we elected to focus on cuts to biomedical research in the form of the proposed cap on indirect costs and realized grant terminations and freezes; additional analyses could examine the potential impacts of further cuts in budget proposals [43], and funding cuts for other agencies (e.g., National Science Foundation, Department of Energy, the Environmental Protection Agency or others). Although the proposed IDC cap could potentially enable greater spending on direct costs of research, the White House’s proposed FY2026 budget included cuts that were far greater than potential reductions from the IDC cap, suggesting that funds would not simply be redistributed from indirect to direct costs. Federal funding cuts to research are likely to have far-reaching and substantial impacts; concerted effort is necessary to understand and quantify their national and international extent [23]. Although the economic multiplier used here has no specified time horizon, we predominantly focus on short-term economic losses [4]. While research funding generates short-term economic benefits [47], cuts will likely lead to long-term health consequences, delays in drug discovery and scientific breakthroughs, disruptions to medical care, lost training opportunities for scientists and physicians, and other downstream consequences that are difficult to quantify and predict, especially in terms of geographic regions [2, 16, 8].

In summary, accounting for commuter flows reveals that cuts to NIH funding will lead to economic losses in many more counties than just the ones that contain research institutions. We project that economic losses exceeding $100,000 from the proposed cap on indirect cost and realized grant terminations in 60% and 22% of counties respectively. Substantial economic impacts of research funding cuts in rural, suburban, and Republican communities are more likely to be underlooked when commuter flows are not taken into consideration. Despite claims that biomedical research cuts are targeted and limited in scope, we find that total economic losses are substantial and geographically widespread, impacting urban, suburban, exurban, and rural communities throughout the U.S.

## Acknowledgments

This work was made possible, in part, by a grant from Coefficient Giving and the Burroughs Wellcome Fund Award 1574584. J.S.W. is supported, in part, by the Simons Foundation award 930382. M.J.H. and J.S.W. are investigators at the University of Maryland-Institute for Health Computing, which is supported by funding from Montgomery County, Maryland and The University of Maryland Strategic Partnership: MPowering the State, a formal collaboration between the University of Maryland, College Park and the University of Maryland, Baltimore. A.H.S. is supported by the Annenberg School for Communication, the Annenberg Public Policy Center, and the Penn Center for Science, Sustainability, and the Media via the Joan Bossert Postdoctoral Research Fellowship. We thank Jeremy Berg, Scott Delaney, Noam Ross, and Ronald Horst for offering significant input on the underlying methodology.

## Data availability

Code and data to conduct analyses are available on Github at doi.org/10.5281/zenodo.22833910. Map visualizations were generated using Esri ArcPro version 3.0.3 [11].

---

**Table S1:**
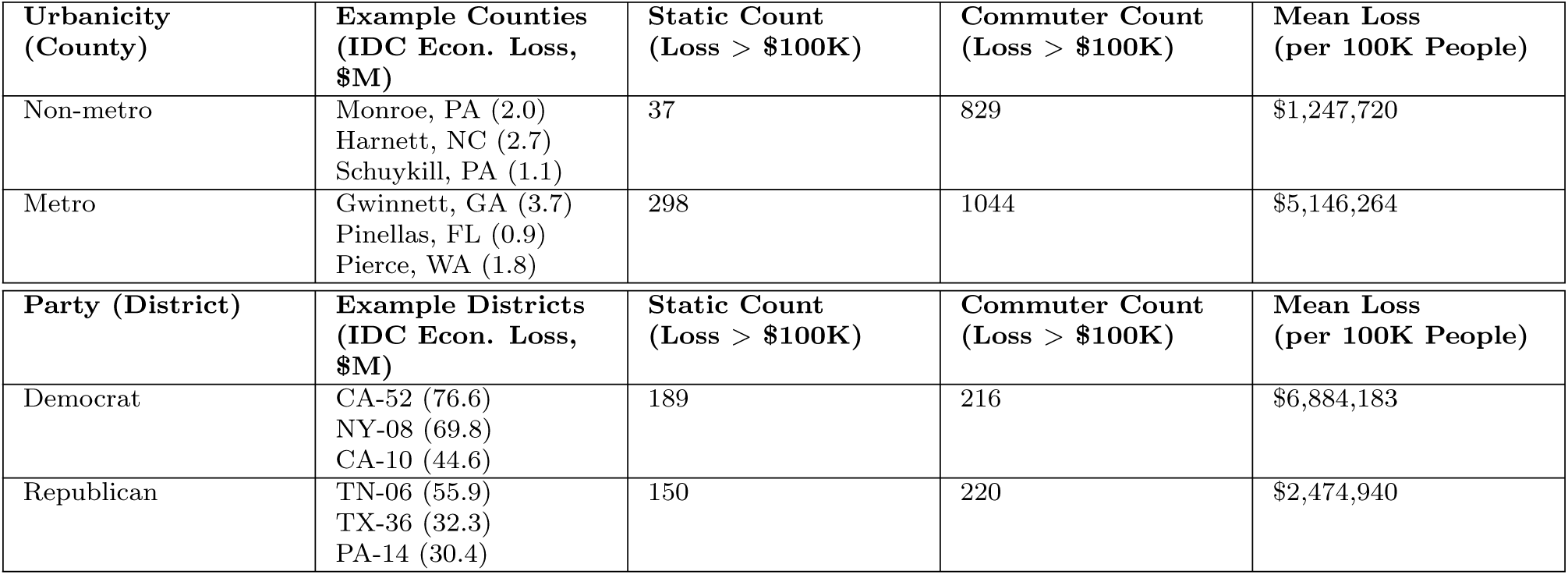
Losses summarized by urbanicity of counties(top) and political party of congressional District (bottom). From left to right, columns indicate the categorization of the county; three example regions with no economic losses from the IDC cap under a static model but over $100K in losses under the commuter model with estimated economic losses in millions of dollars in parentheses (counties selected based on largest population sizes; districts selected based on greatest losses); the number of regions with over $100K in losses under the static model; the number of regions with over $100K in losses under the commuter model; and total economic losses per 100,000 population.

| Urbanicity<br>(County) | Example Counties<br>(IDC Econ. Loss,<br>\$M) | Static Count<br>(Loss > \$100K) | Commuter Count<br>(Loss > \$100K) | Mean Loss<br>(per 100K People) |
| --- | --- | --- | --- | --- |
| Non-metro | Monroe, PA (2.0)<br>Harnett, NC (2.7)<br>Schuylkill, PA (1.1) | 37 | 829 | \$1,247,720 |
| Metro | Gwinnett, GA (3.7)<br>Pinellas, FL (0.9)<br>Pierce, WA (1.8) | 298 | 1044 | \$5,146,264 |
| Party (District) | Example Districts<br>(IDC Econ. Loss,<br>\$M) | Static Count<br>(Loss > \$100K) | Commuter Count<br>(Loss > \$100K) | Mean Loss<br>(per 100K People) |
| Democrat | CA-52 (76.6)<br>NY-08 (69.8)<br>CA-10 (44.6) | 189 | 216 | \$6,884,183 |
| Republican | TN-06 (55.9)<br>TX-36 (32.3)<br>PA-14 (30.4) | 150 | 220 | \$2,474,940 |

**Table S2:** Losses summarized by American Communities Project county typology. From left to right, columns indicate: community name; total population of residents in counties of a given type; number of counties of a given type; count (and percentage) of counties with economic losses from the proposed IDC cap exceeding $100K under a static model; count (and percentage) of counties with economic losses from the proposed IDC cap exceeding $100K under a commuter model; mean economic losses from the proposed IDC cap per 100,000 across the given type; count (and percentage) of counties with economic losses from terminated/frozen grants exceeding $100K under a static model; count (and percentage) of counties with economic losses from terminated/frozen grants exceeding $100K under a commuter model; mean economic losses from terminated/frozen grants per 100,000 across the given type.

| Name | Pop. | Count | Static IDC | Comm. IDC | Mean IDC Loss<br>(per 100K pop.) | Static Term. | Comm. Term. | Mean Term. Loss<br>(per 100K pop.) |
| --- | --- | --- | --- | --- | --- | --- | --- | --- |
| Big Cities | 83,134,988 | 48 | 47 (98%) | 48 (100%) | \$7,820,948 | 38 (79%) | 45 (94%) | \$641,948 |
| Urban Burbs | 71,469,895 | 109 | 69 (63%) | 108 (99%) | \$5,480,592 | 20 (18%) | 101 (93%) | \$470,003 |
| Exurbs | 33,981,138 | 194 | 39 (20%) | 179 (92%) | \$3,396,376 | 3 (2%) | 99 (51%) | \$285,572 |
| College Towns | 25,859,484 | 172 | 102 (59%) | 158 (92%) | \$6,956,722 | 38 (22%) | 73 (42%) | \$1,778,671 |
| Rural Middle America | 24,398,585 | 627 | 10 (2%) | 443 (71%) | \$1,608,434 | 0 (0%) | 78 (12%) | \$137,558 |
| Hispanic Centers | 18,116,081 | 178 | 10 (6%) | 73 (41%) | \$1,107,801 | 2 (1%) | 21 (12%) | \$104,686 |
| Graying America | 17,135,196 | 395 | 11 (3%) | 203 (51%) | \$1,328,033 | 2 (1%) | 30 (8%) | \$72,291 |
| African American South | 12,907,939 | 272 | 17 (6%) | 157 (58%) | \$2,799,827 | 11 (4%) | 59 (22%) | \$646,627 |
| Middle Suburbs | 12,475,573 | 57 | 4 (7%) | 57 (100%) | \$2,065,752 | 2 (4%) | 42 (74%) | \$199,237 |
| Working Class Country | 11,002,531 | 280 | 3 (1%) | 169 (60%) | \$1,356,176 | 0 (0%) | 62 (22%) | \$539,751 |
| Evangelical Hubs | 10,040,423 | 375 | 3 (1%) | 169 (45%) | \$836,710 | 0 (0%) | 32 (9%) | \$217,701 |
| Military Posts | 10,030,094 | 76 | 9 (12%) | 63 (83%) | \$1,238,554 | 2 (3%) | 23 (30%) | \$179,919 |
| LDS Enclaves | 3,929,965 | 39 | 4 (10%) | 18 (46%) | \$3,468,810 | 1 (3%) | 4 (10%) | \$103,889 |
| Aging Farmlands | 1,107,544 | 268 | 0 (0%) | 11 (4%) | \$499,234 | 0 (0%) | 0 (0%) | \$30,537 |
| Native American Lands | 846,402 | 44 | 1 (2%) | 8 (18%) | \$426,829 | 1 (2%) | 2 (5%) | \$313,577 |

**Figure S1:**
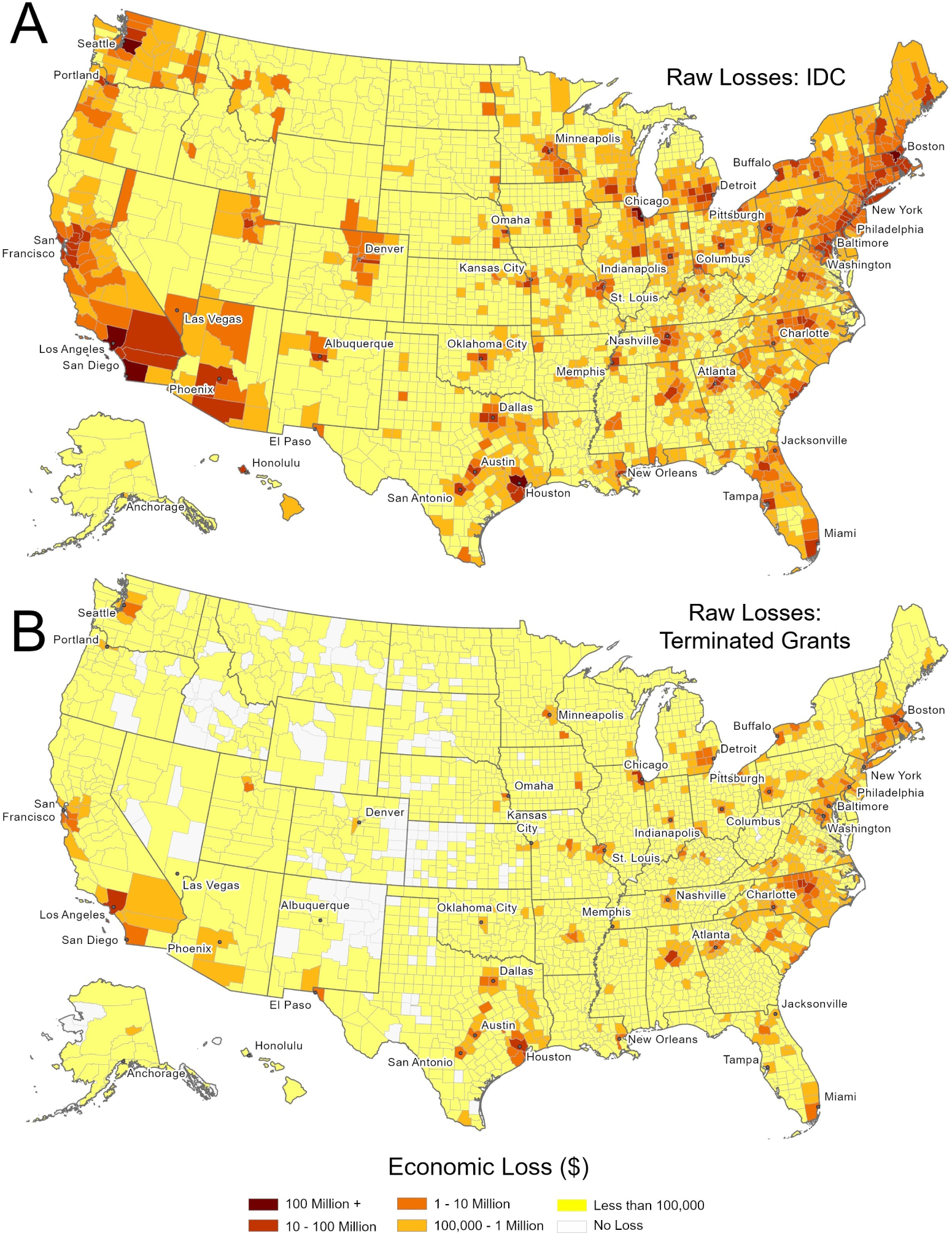
Raw (unmultiplied) losses at the county level. Panel A shows losses from IDC cap, and Panel B shows losses from frozen/terminated grants. County-level raw values are available and archived on Zenodo at doi.org/10.5281/zenodo.22833910.

**Figure S2:**
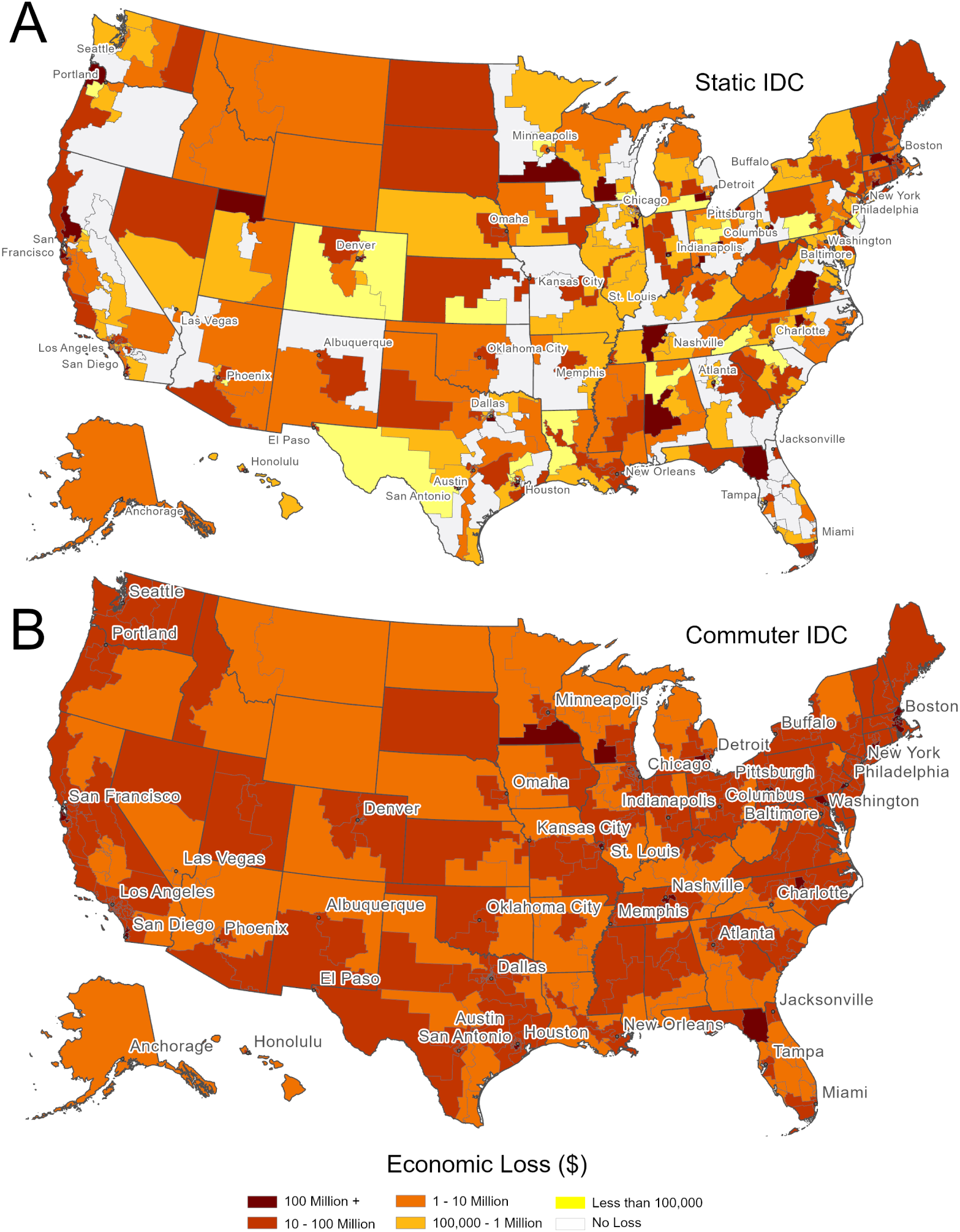
Economic losses for IDC cap by congressional district (119th Congress). Panel A shows estimates when using a static method, and Panel B shows estimates when using commuter flows (in USD). District-level values are available and archived on Zenodo at doi.org/10.5281/zenodo.22833910.

**Figure S3:**
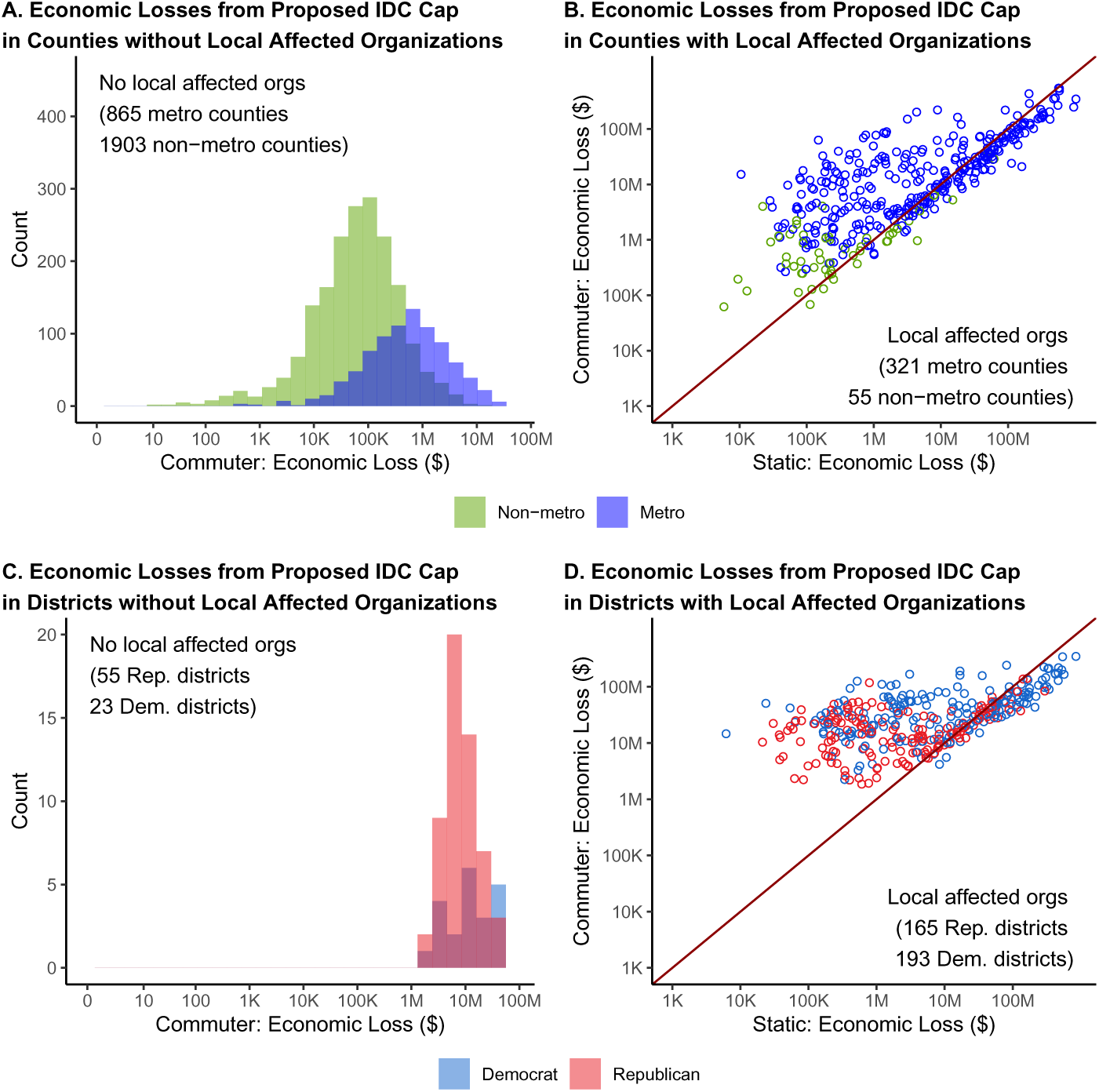
Comparison of economic losses from IDC cap across static versus commuter models depending on urbanicity (top) or partisanship (bottom). Urbanicity is split into non-metro (green) or metro (blue). Partisanship is split into Democratic (blue) or Republican (red). Comparisons based on partisanship are estimated at the congressional district level. Regions are split into distinct panels depending on whether an individual region lacks (left, A and C) or contains (right, B and D) local affected organizations, as in Figure 3.

**Figure S4:**
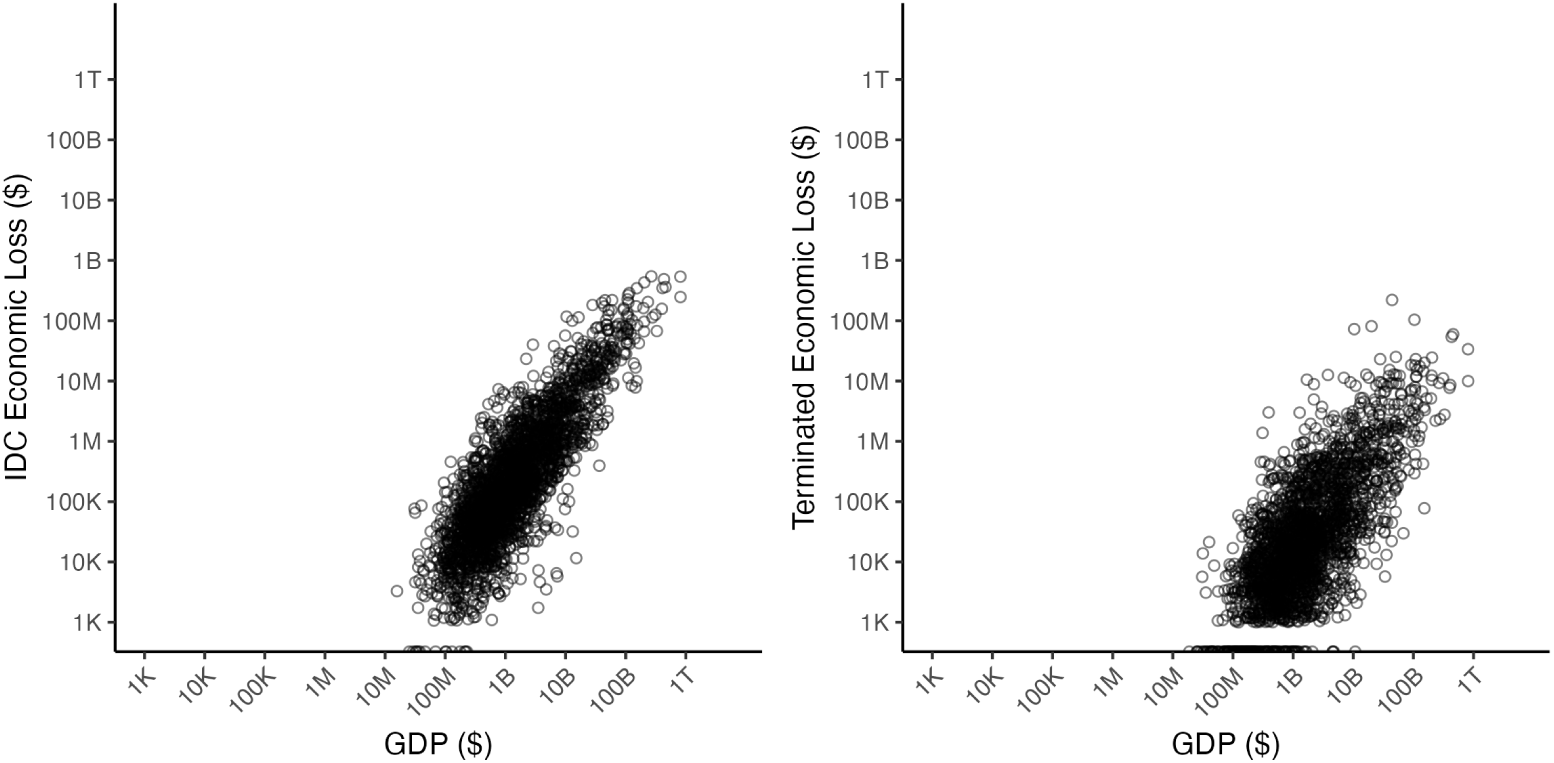
Anticipated economic losses are associated with GDP at the county level. Log-scaled GDP for 2024 is compared to economic losses from the proposed IDC cap (left) or terminated grants (right).

**Figure S5:**
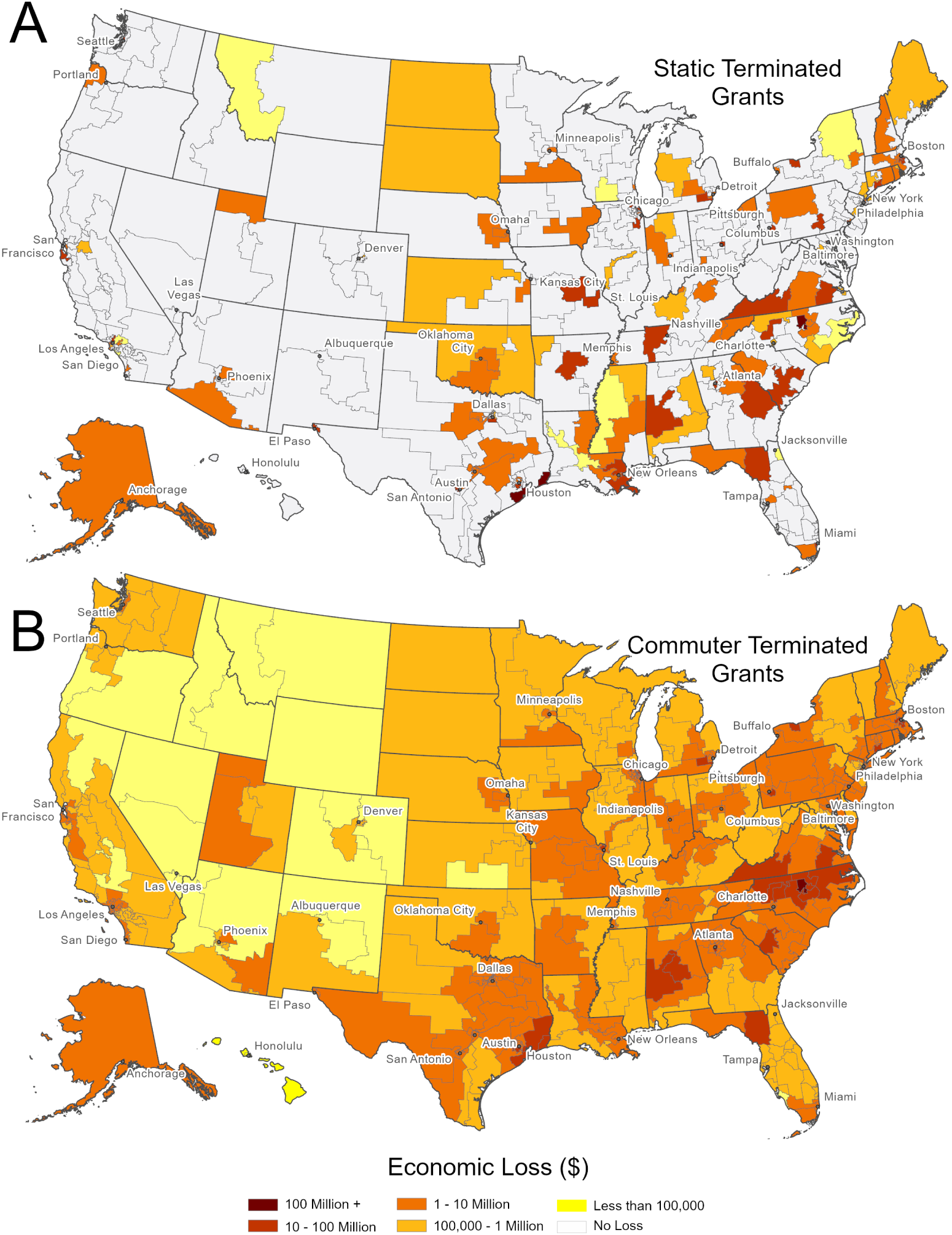
Economic losses from terminated grants by congressional district (119th Congress). Panel A shows estimates when using a static method, and Panel B shows estimates when using commuter flows (in USD). District-level values are available and archived on Zenodo at doi.org/10.5281/zenodo.22833910.

**Figure S6:**
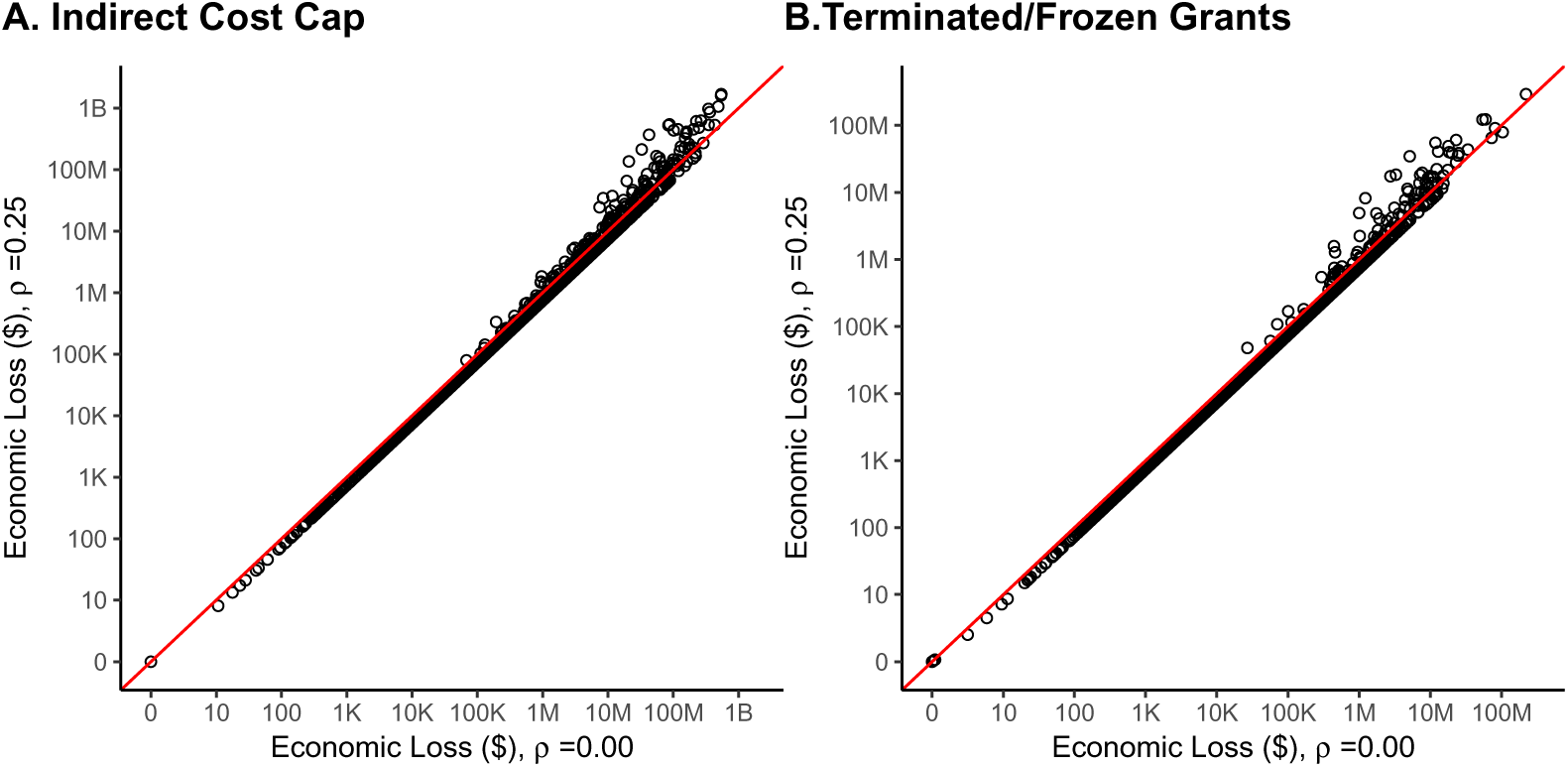
The estimated county-level economic loss under the commuter model (x-axis, *ρ* = 0.00) is well-aligned with estimates where a quarter of funding is static and the rest follows commuter flows (y-axis, *ρ* = 0.25), (*R*^2^ = 0.83 and *R*^2^ = 0.90 for the IDC cap and grant terminations/freezes respectively). Values correspond to county-level economic losses from the proposed IDC cap (left) or grant terminations/freezes (right). The red line indicates perfect alignment (y=x). Values are log-scaled.

**Figure S7:**
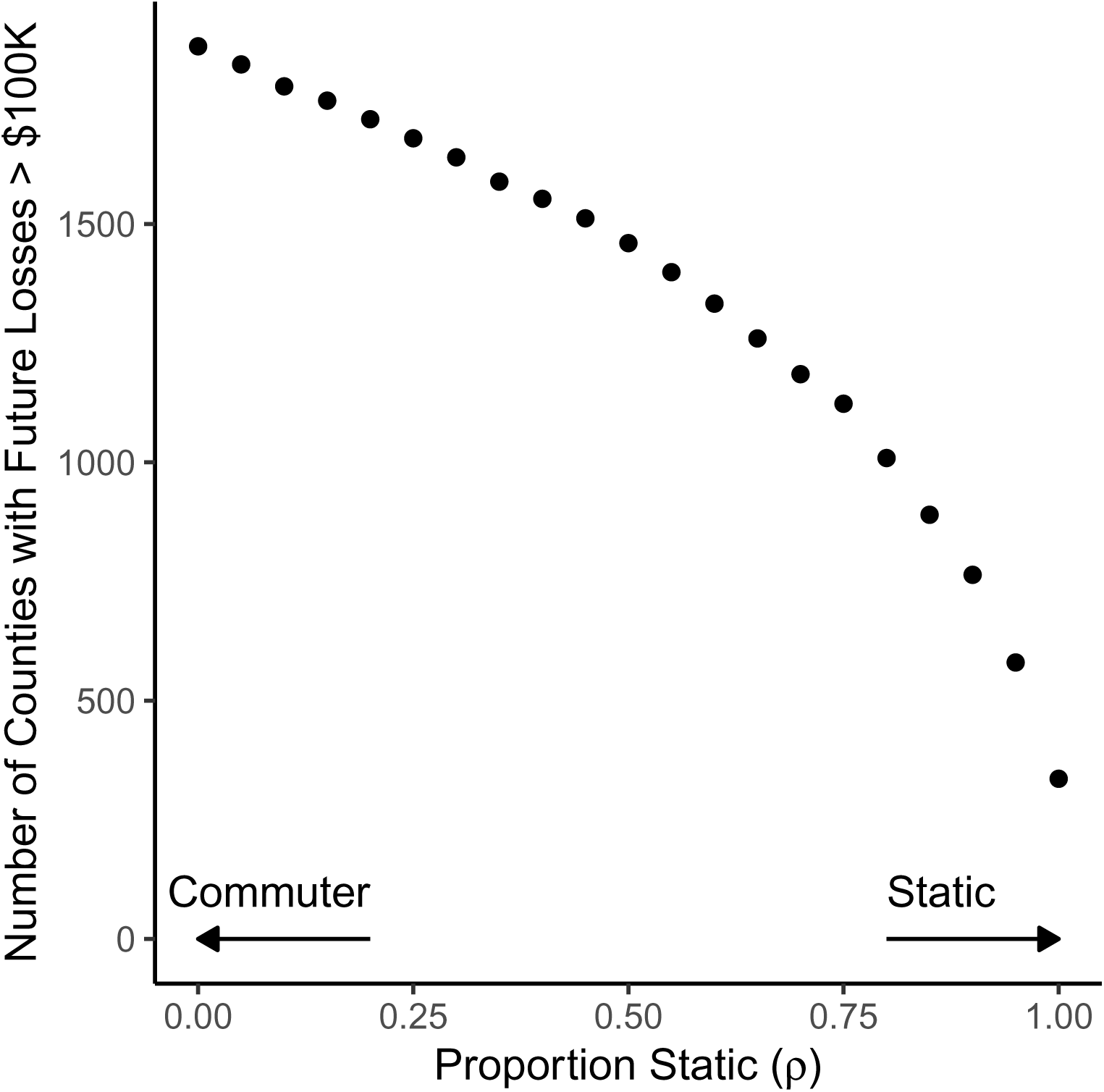
The number of counties with greater than $100,000 in economic losses from the IDC cap (y-axis) when the share of funding that remains static. (*ρ*) **(x-axis) varies**. Values of *ρ* vary from 0 (commuter model) to 1 (static model).

**Figure S8:**
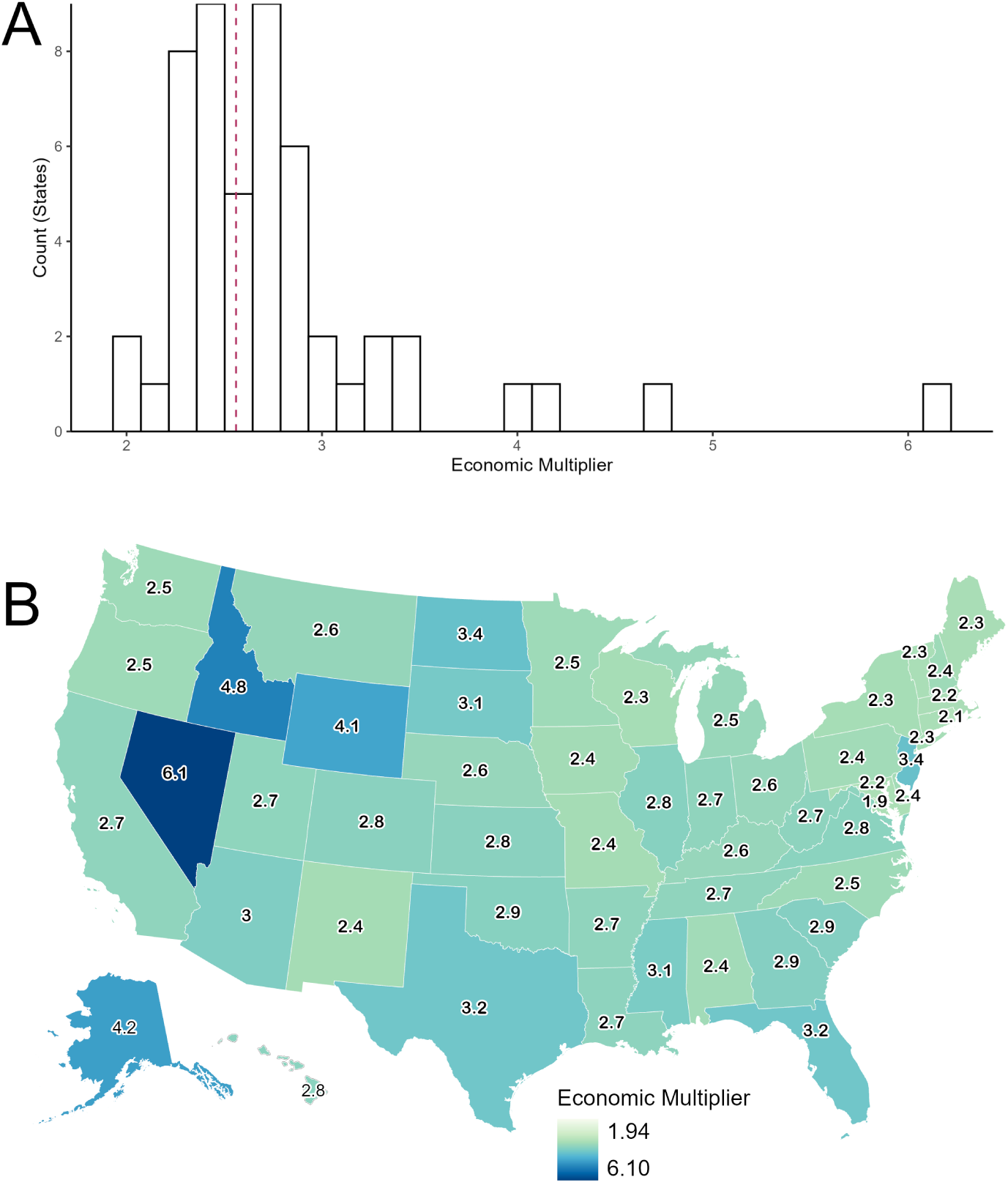
The economic multiplier for NIH investment varies across states. Variation is displayed as: (A) a histogram where the national value (2.56) is indicated as a dashed maroon line and (B) a map where states are shaded according to their economic multipliers, which is also displayed as an annotation. The economic multiplier varies across states from 1.94 in Maryland to 6.10 in Nevada.

**Figure S9:**
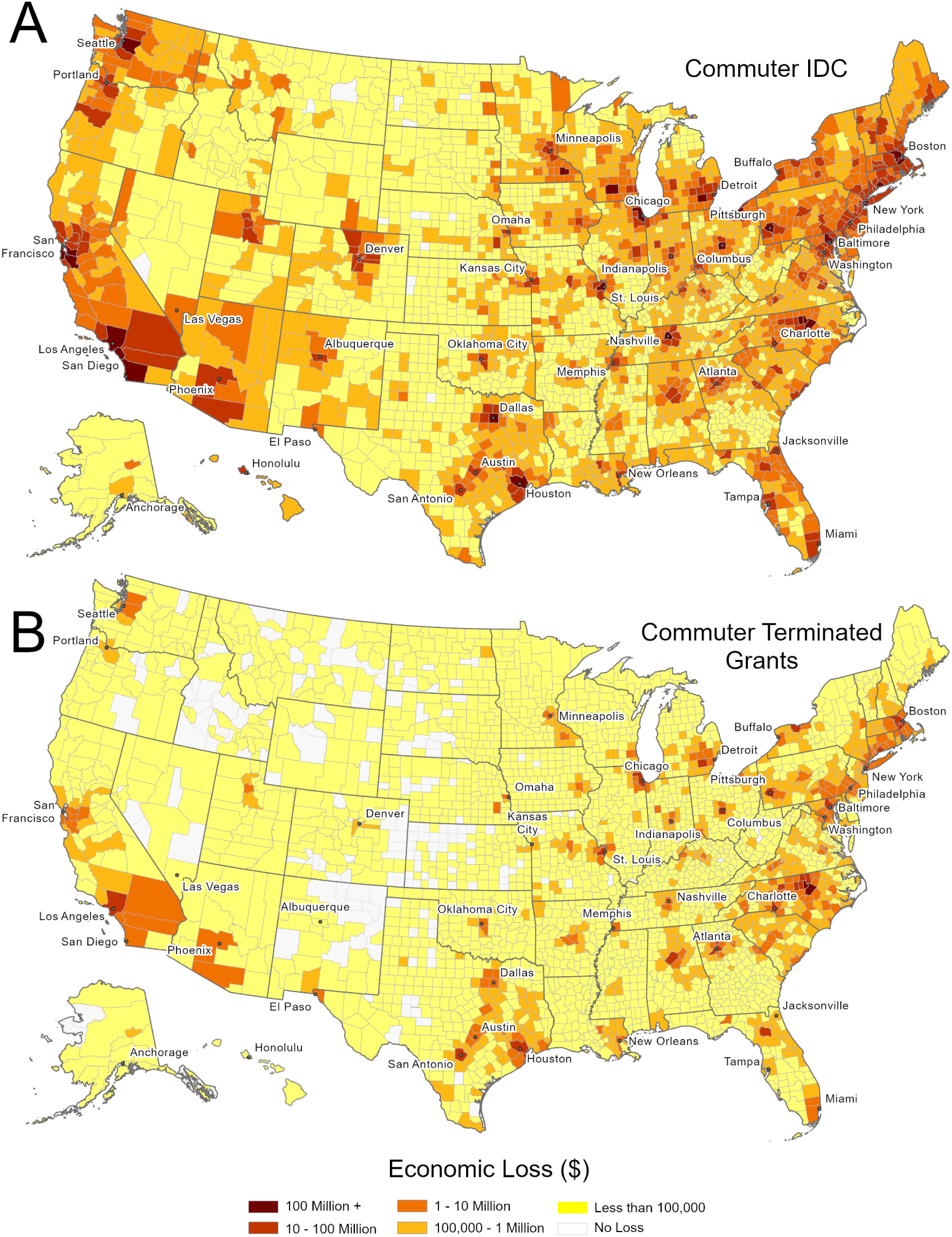
Estimated economic losses are robust to using a national economic multiplier. Economic losses are displayed at the county level. Panel A shows losses from IDC cap, and Panel B shows losses from frozen/terminated grants. County-level values estimated using a national multiplier are available and archived on Zenodo at doi.org/10.5281/zenodo.22833910.

**Figure S10:**
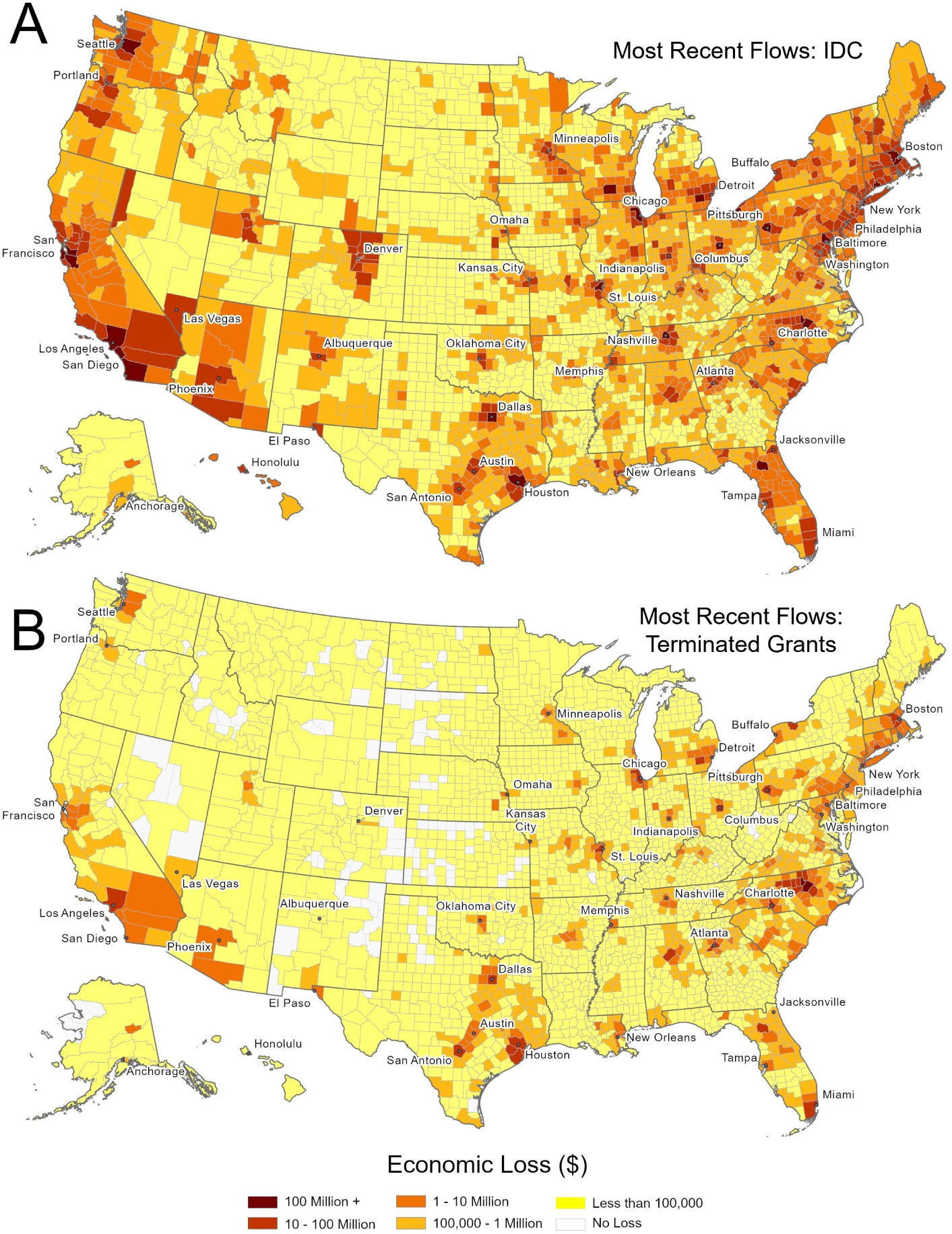
Estimated economic losses are robust to using the most recent available commuter flows. Economic losses are displayed at the county level. Losses are estimated using flows for 2023 for all states except for Michigan and Alaska, where the most recent available commuter flows were from 2022 and 2016 respectively. Panel A shows losses from IDC cap, and Panel B shows losses from frozen/terminated grants. County-level values estimated using the most recent available commuter flows are available and archived on Zenodo at

**Figure S11:**
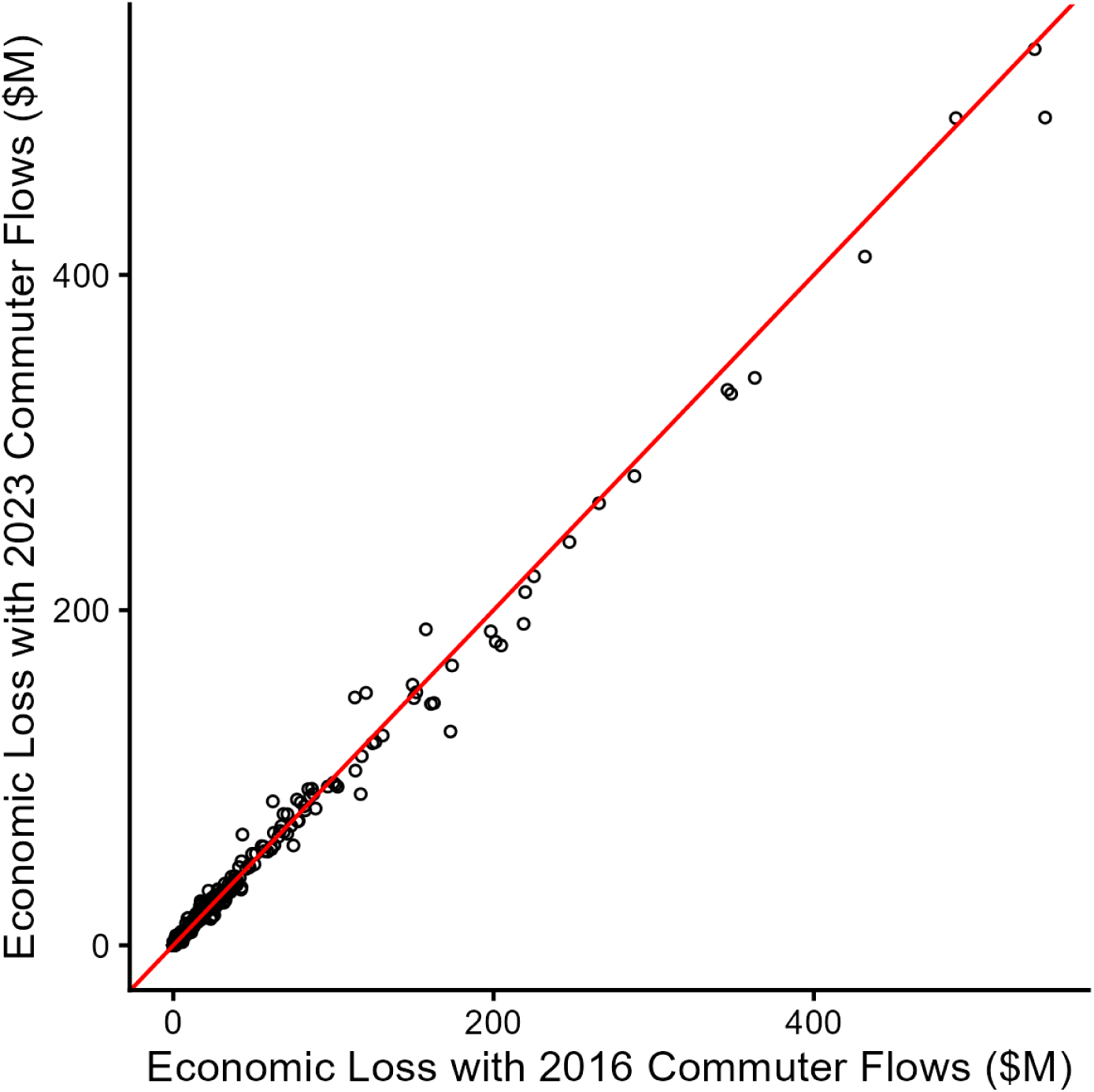
The estimated economic loss using 2016 commuter flows (x-axis) is well-aligned with estimates using the most recent available commuter flows (y-axis), (*R*^2^ = 0.99, Spearman’s *ρ* = 0.94). Values correspond to county-level economic losses from the proposed IDC cap. Losses are reported in millions of dollars. The red line indicates perfect alignment (y=x). States without available commuter flows for 2023 (Michigan and Alaska) are excluded.

**Figure S12:**
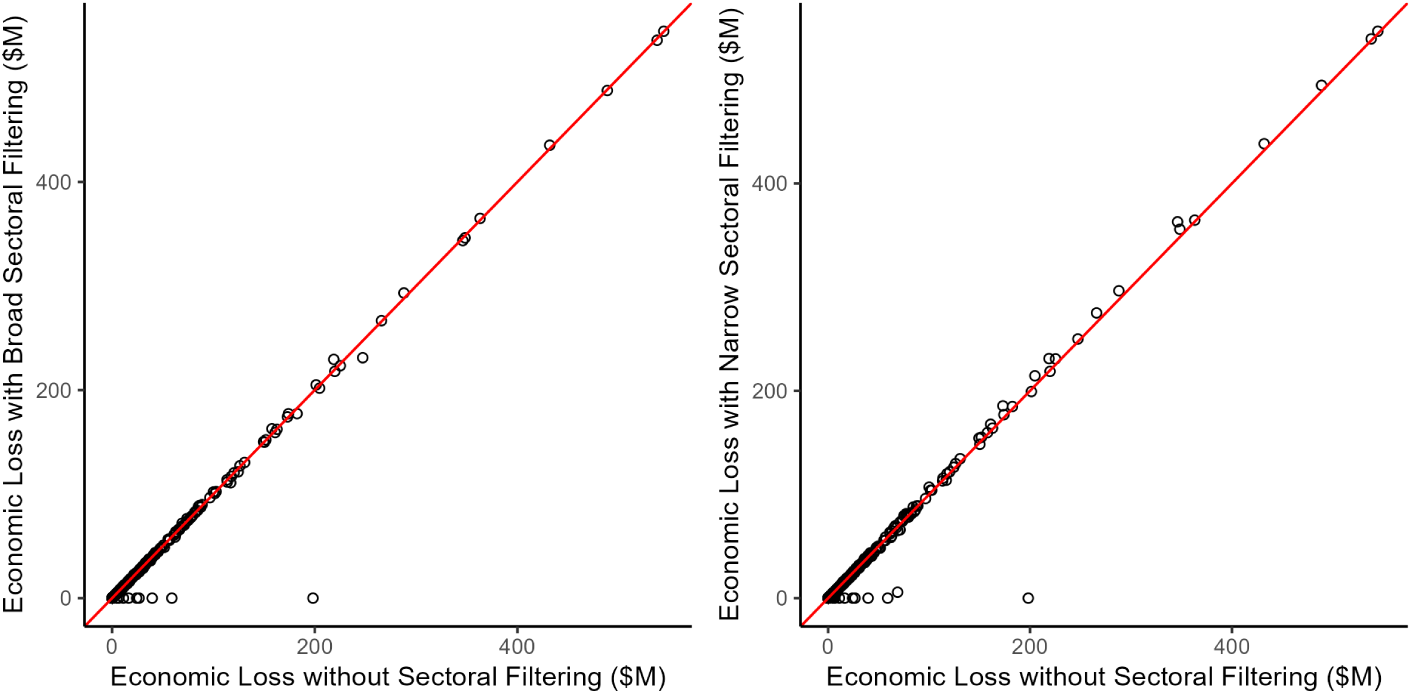
Estimated economic losses are robust to sectoral filtering (*R*^2^ = 0.98 for both broad and narrow sectoral filtering). Values correspond to county-level economic losses from the proposed IDC cap without (x-axis) or with (y-axis) sectoral filtering. We compare a broad set of relevant sectors (left) to a more narrow set of relevant sectors (right). Losses are reported in millions of dollars. The red line indicates perfect alignment (y=x).

**Figure S13:**
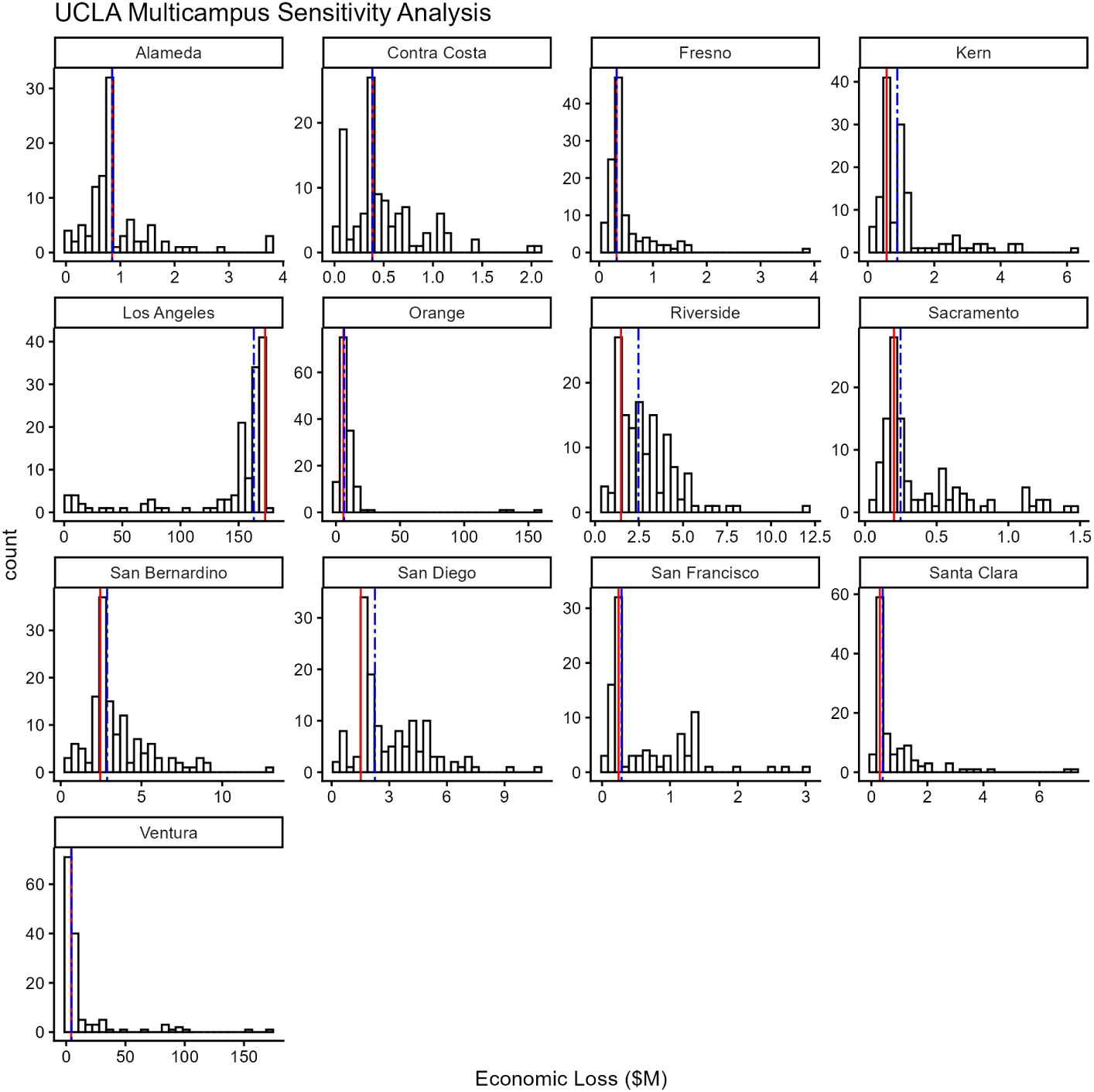
Estimated economic losses from UCLA to connected counties in multicampus sensitivity analysis. Panels display counties connected to UCLA, defined as those with at least ten residents who commute to the census tract containing UCLA’s main campus. Each panel illustrates the estimated economic loss from the proposed IDC cap (in millions of dollars), seeding losses in each of the 329 UCLA-affiliated locations. The red solid line indicates the main model specification, when losses are geolocated to the census tract where UCLA’s main campus is located. The blue dashed line indicates the median estimated losses across scenarios.

**Figure S14:**
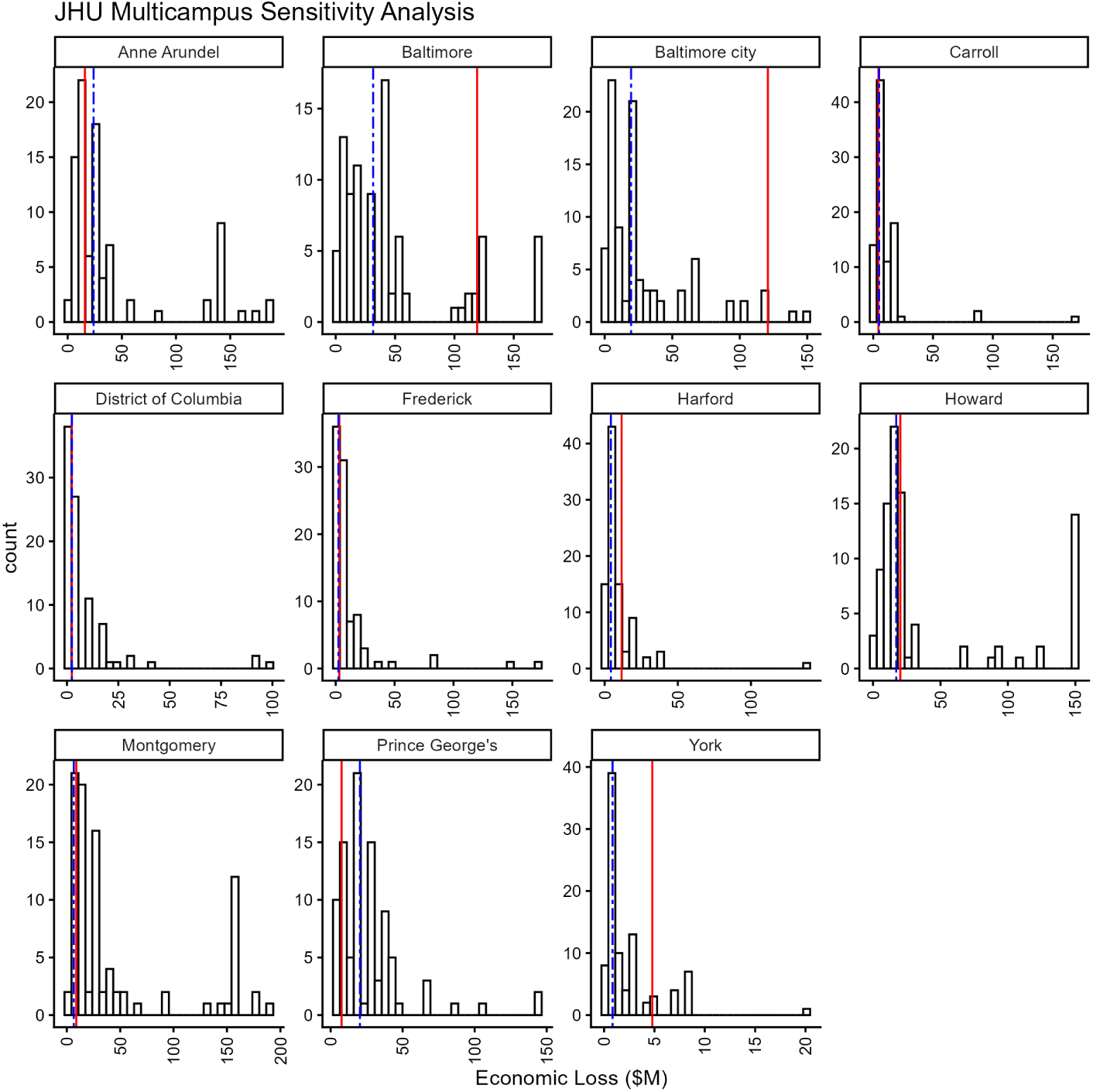
Estimated economic losses from Johns Hopkins to connected counties in multicampus sensitivity analysis. Panels display counties connected to Johns Hopkins, defined as those with at least ten residents who commute to the census tract containing Johns Hopkins’ main campus. Each panel illustrates the estimated economic loss from the proposed IDC cap (in millions of dollars), seeding losses in each of the 91 Johns Hopkins-affiliated locations. The red solid line indicates the main model specification, when losses are geolocated to the census tract where Johns Hopkins’ main campus is located. The blue dashed line indicates the median estimated losses across scenarios.

## Supplemental File: Census Tract Assignment

### Introduction

We used case studies in Connecticut, Massachusetts, New York, West Virginia, and Wisconsin for our analysis as these states had public parcel data that were findable and usable by the public. Below are the reports on *points whose intersecting parcels did not fully fall into one Census tract*. We do not count parcels that were not 100% contained in a tract due to small sliver polygons. Sliver polygons occur when boundaries do not fully align geometrically due to line generalization, heads up digitizing, or coastal/road digitizing issues, etc.

### Method

The analysis was conducted using ArcGIS Pro. We retrieved the polygonal parcels that these institutions fell into using a select by location function. There are often more parcels than points because apartment buildings and some multi-floor buildings have overlapping polygons (i.e., polygons are stacked on top of each other). We then measured which percent of the point’s parcel fell into the same tract that the point was assigned to.

For Connecticut and New York, we removed parcels that were road grids with no buildings (these were polygonised road systems). If a point did not overlay a parcel (e.g., it was in between two parcels, most likely the road), it was snapped to the edge of its closest parcel. We used the Identity tool to slice parcels into sections based on tract boundaries. Then, we dissolved parcels by parcel ID and through min, max, count and sum summary statistics, we found which percentage of the parcel was in the tract that the point had been assigned to.

We define **incorrect assignment** as a condition where the address point is located in a parcel where less than 50% of its area falls inside the tract that the point was assigned to.

### Summary

Out of 566 institutions tested, we identified two points (*n*=2**) that fell into incorrect tracts (i.e., 564 of 566 are correctly assigned).** These were distributed across the four states: CT 41 institutions (1 wrong case), MA 172 institutions (0 wrong cases), NY 174 institutions (0 wrong cases), WV 28 institutions (0 wrong cases), WI 151 institutions (1 wrong case). Due to incompleteness of the New York State data, 21 additional institutions could not be validated. **The incorrectly placed institutions are University of Hartford and University of Wisconsin-Parkside.**

## STATE DETAILS

### Connecticut

There were 41 point-level institutions in Connecticut. One was assigned to the incorrect tract.

Two (2) parcels were not fully contained by census tracts. One address, where only 3% of its parcel’s area was within its assigned tract, was incorrectly assigned. *Parcel Data Source: Connecticut CAMA and Parcel Layer (2024-2026) accessed via geodata.ct.gov/datasets*.

| Institution Name | Parcel Owner | Percentage of Parcel Area within Point's Assigned Tract |
| --- | --- | --- |
| University of Hartford | WATKINSON SCHOOL | 3.962 |
| GYNION, LLC | SHIKHMAN OLEG & VICTORIA | 61.591 |

### Massachusetts

There were 172 point-level institutions in Massachusetts. These points occupied 163 different parcels (max points in a parcel = 6). All (100%) points were assigned to the tract where their parcel had a majority overlap with the tract. *Parcel Data Source: MassGIS Data: Property Tax Parcels (June 2026) accessed via mass.gov/info-details/massgis-data-property-tax-parcels, by filling out a webform and a geodatabase was sent via email*.

| **Institution Name** | **Parcel Owner** | **Percentage of**<br>**Parcel Area**<br>**within Point's**<br>**Assigned**<br>**Tract** |
| --- | --- | --- |
| XGENOMES CORP. | MARLEY JAH-NESTA | 65.388 |
| AQUINNAH<br>PHARMACEUTICALS,<br>INC. | WOLOZIN BENJAMIN | 78.190 |
| AMPLIFIDX INC | 8-9 MONUMENT LLC | 78.793 |
| BLOOMER HEALTH<br>TECH INC | CURRAN, THOMAS | 80.714 |
| IBIS REPRODUCTIVE<br>HEALTH | LOMBARDO, PETER J. | 89.248 |

### New York

There were 195 point-level institutions in New York, however because the New York State parcel dataset was not complete, only 174 points fell within the NY parcel boundary. The points were in 219 parcels (due to parcel polygon overlaps). Four parcels were not contained by a tract. However, all address points were assigned to the tract where their parcel had a majority overlap with the tract. *Parcel Data Source: NYS_Tax_Parcels_Public (FeatureServer) (May 2026) accessed via gisservices.its.ny.gov*.

| Institution Name | Parcel Owner | Percentage of Parcel Area within Point's Assigned Tract |
| --- | --- | --- |
| NEW YORK UNIVERSITY | NEW YORK UNIVERSITY | 61.264 |
| RENSSELAER POLYTECHNIC INSTITUTE | R P I | 71.887 |
| STATE UNIVERSITY OF NEW YORK AT ALBANY | State of New York | 79.603 |
| NEW YORK STRUCTURAL BIOLOGY CENTER and ADVANCED SCIENCE RESEARCH CENTER | CITY COLLEGE | 85.228 |

### West Virginia

There were 28 point-level institutions in West Virginia; 0 points were assigned to the incorrect tract. There were 27 parcels. All 28 organizations fell into parcel polygons that were 100% contained in their respective census tracts, thus we provide no table below. *Parcel Data Source: WV Department of Tax and Revenue Tax Maps - Surface and Mineral Parcels (Statewide) (2025) accessed via wvgis.wvu.edu/data*.

### Wisconsin

There were 151 point-level institutions in Wisconsin; 1 point was assigned to the incorrect tract. There were 858 parcels. Two cases have the majority of the parcel in the tract we assigned the point to (61% and 74% of the parcel is in the tract), and one point only had 18% of its parcel in the tract, indicating that one point was potentially misassigned to the wrong tract. *Parcel Data Source: Wisconsin Statewide Parcel Layer Version 11.0.0 (2025) accessed via doa.wi.gov*.

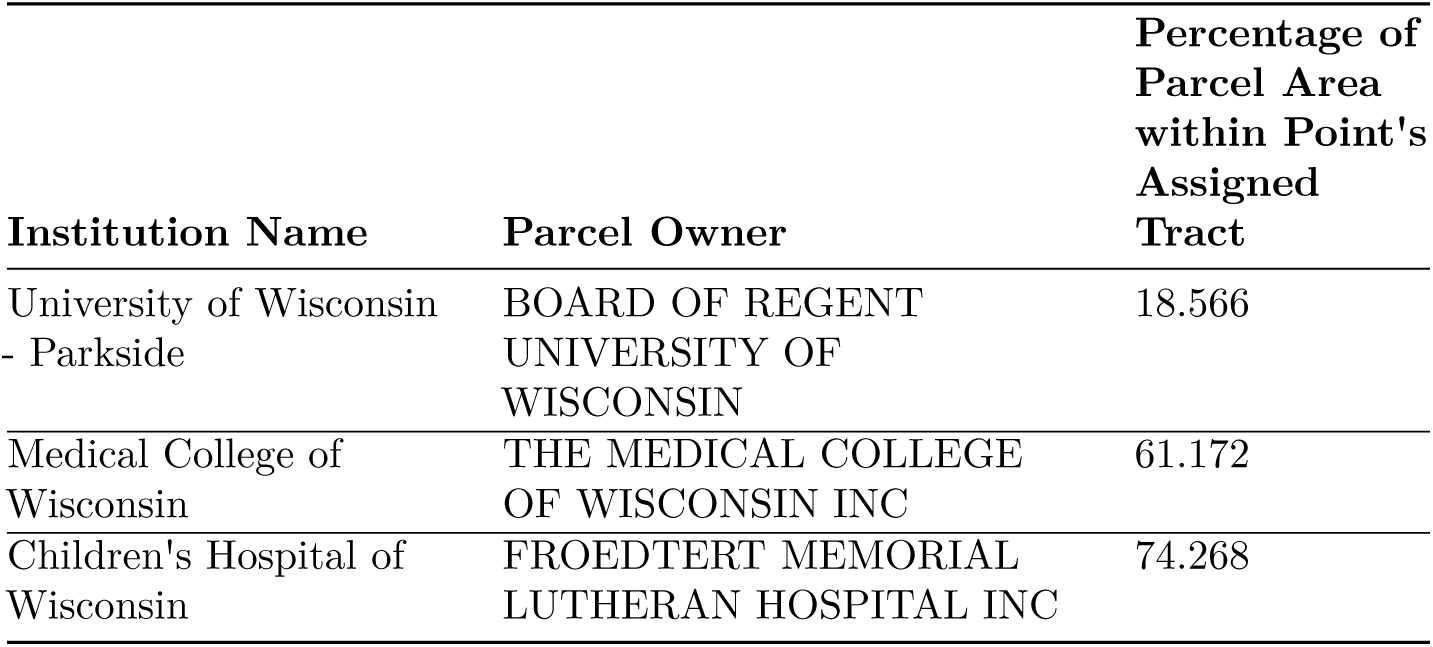

## GRAPHICAL EXAMPLES

The blue area is a single parcel for the Masonic Medical Research Laboratory in Wisconsin, and the black outline is a census tract.

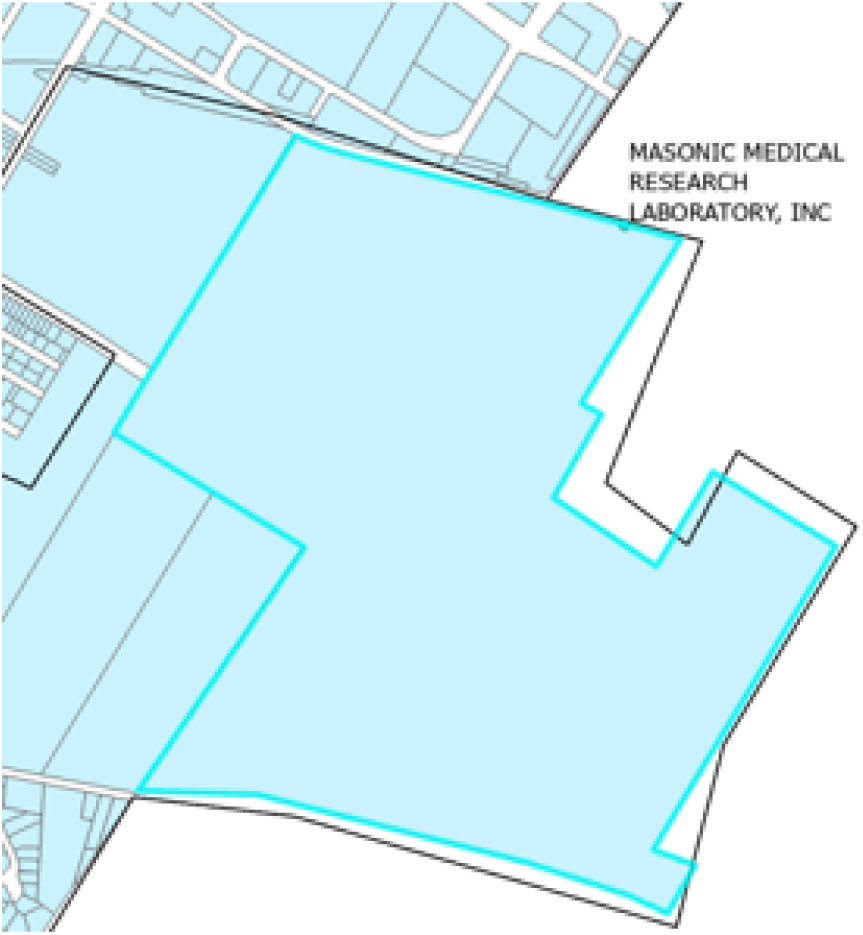

The University of Wisconsin-Parkside was associated with a Census tract that did not contain the majority of the university’s parcel (in grey). The orange polygon represents a bounding box.

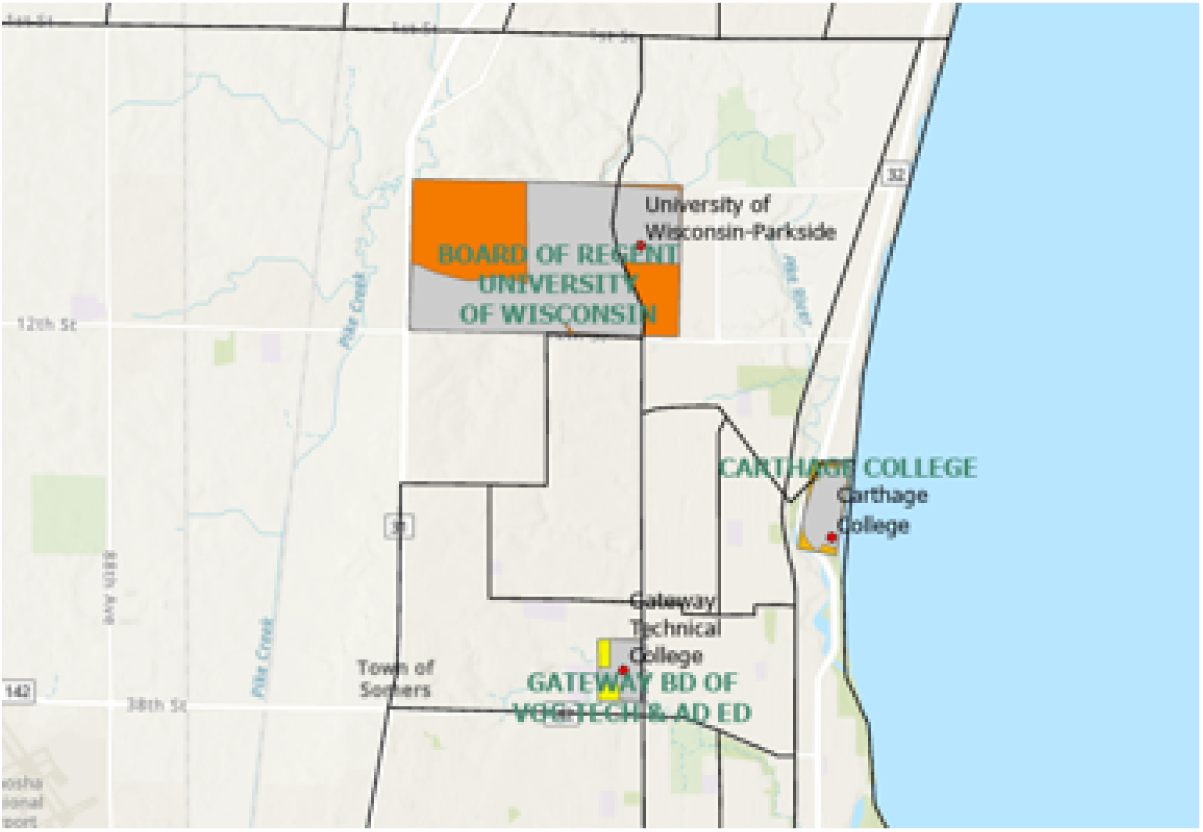

The University of Hartford point was assigned to a Census tract (i.e., west of the black line) that did not contain much of the University’s campus, but did contain other parcels owned by the University of Hartford (in all caps).

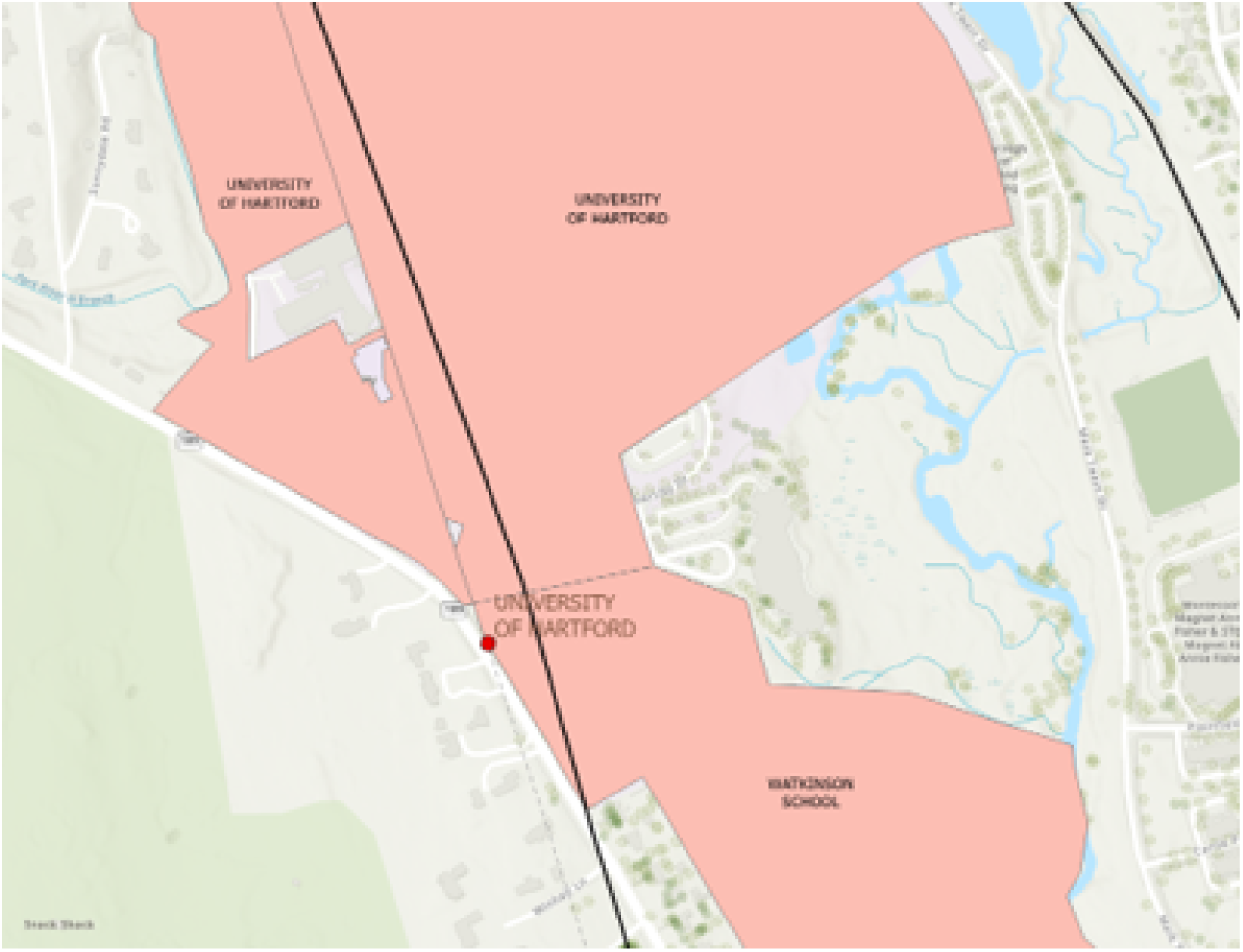

